# Disrupted Brain Connectivity in Children Treated with Therapeutic Hypothermia for Neonatal Encephalopathy

**DOI:** 10.1101/2020.09.23.20200014

**Authors:** Arthur P.C. Spencer, Jonathan C.W. Brooks, Naoki Masuda, Hollie Byrne, Richard Lee-Kelland, Sally Jary, Marianne Thoresen, James Tonks, Marc Goodfellowi, Frances M. Cowan, Ela Chakkarapani

**Affiliations:** Clinical Research and Imaging Centre, University of Bristol, Bristol, UK; School of Psychological Science, University of Bristol, Bristol, UK; Department of Mathematics, State University of New York at Buffalo, Buffalo, NY, USA; Computational and Data-Enabled Science and Engineering Program, State University of New York at Buffalo, Buffalo, NY, USA; Translational Health Sciences, Bristol Medical School, University of Bristol, Bristol, UK; Faculty of Medicine, Institute of Basic Medical Sciences, University of Oslo, Oslo, Norway; Neonatal intensive care unit, St Michael’s Hospital, University Hospitals Bristol and Weston NHS Foundation Trust, Bristol, UK; University of Exeter Medical School, Exeter, UK; Living Systems Institute, University of Exeter, Exeter, UK; Wellcome Trust Centre for Biomedical Modelling and Analysis, University of Exeter, Exeter, UK; EPSRC Centre for Predictive Modelling in Healthcare, University of Exeter, Exeter, UK; College of Engineering, Mathematics and Physical Sciences, University of Exeter, Exeter, UK; Department of Paediatrics, Imperial College London, London, UK

**Keywords:** Neonatal encephalopathy, therapeutic hypothermia, white matter, structural connectivity, brain networks, diffusion-weighted imaging

## Abstract

Therapeutic hypothermia following neonatal encephalopathy due to birth asphyxia reduces death and cerebral palsy. However, school-age children without cerebral palsy treated with therapeutic hypothermia for neonatal encephalopathy still have reduced performance on cognitive and motor tests, attention difficulties, slower reaction times and reduced visuo-spatial processing abilities compared to typically developing controls. We acquired diffusion-weighted imaging data from school-age children without cerebral palsy treated with therapeutic hypothermia for neonatal encephalopathy at birth, and a matched control group. Voxelwise comparison of fractional anisotropy (33 cases, 36 controls) confirmed microstructural alterations in widespread areas of white matter in cases, particularly in the fornix, corpus callosum, anterior and posterior limbs of the internal capsule bilaterally and cingulum bilaterally. In structural brain networks constructed using probabilistic tractography (22 cases, 32 controls), graph-theoretic measures of strength, local and global efficiency, clustering coefficient and characteristic path length were found to correlate with IQ in cases but not controls. Network-based statistic analysis implicated brain regions involved in visuo-spatial processing and attention, aligning with previous behavioural findings. These included the precuneus, thalamus, left superior parietal gyrus and left inferior temporal gyrus. Our findings demonstrate that, despite the manifest successes of therapeutic hypothermia, brain development is impaired in these children.

## 1 Introduction

Neonatal encephalopathy (NE), which often results from perinatal asphyxia, leads to a high risk of death or disability, including cerebral palsy (CP) (Azzopardi et al., 2014; Marlow, 2005; O’Connor et al., 2017; Robertson et al., 1989). In the UK, approximately 2.6 per 1000 live births in 2015 were affected by NE secondary to perinatal asphyxia (Gale et al., 2018). The recommended treatment for NE (National Institute for Clinical Excellence (NICE), 2010) is therapeutic hypothermia (TH), which consists of reducing the infant’s core temperature to 33.5°C for three days, commencing as soon as possible after the asphyxia (Azzopardi et al., 2009; Rutherford et al., 2010). TH reduces the chance of death and disability at 18 months (Jacobs et al., 2013), reduces likelihood and severity of CP (Jary et al., 2015) and increases the incidence of survival with an IQ > 85 (Azzopardi et al., 2014). However, recent studies have shown that children aged 6-8 years, who underwent TH at birth for NE and did not develop CP, perform worse in motor and cognitive tests than controls (Jary et al., 2019; Lee-Kelland et al., 2020) and have attention difficulties, slower reaction times and reduced visuo-spatial processing abilities (Tonks et al., 2019). These motor deficits are not predicted by 18-month developmental scores (Jary et al., 2019). Thus, despite the reduced occurrence of severe disabilities following TH, aspects of brain development remain affected by NE.

Studies on children born with NE, prior to widespread use of TH (Gao et al., 2012; Ly et al., 2015; Martinez-Biarge et al., 2012), and on animal models (Chakkarapani et al., 2010; Kyng et al., 2015; Yue et al., 1997) indicate damage to white matter and subcortical structures, caused by hypoxic-ischaemic brain injury. Some studies have shown an association between hypothermia/rewarming and subcortical white matter apoptosis independent of hypoxic-ischemic brain injury (O’Brien et al., 2019; Wang et al., 2016), whereas other findings suggest no impact of hypothermia on the subcortical white matter (Gressens et al., 2008). It is unknown how the interplay between the damage mechanisms of NE and the effects of TH impact brain development.

Diffusion-weighted imaging (DWI) provides a non-invasive tool for investigating white matter microstructure. Measurement of diffusion of water molecules through brain tissue allows calculation of diffusion metrics such as fractional anisotropy (FA), which is related to its microstructural properties. FA is affected by properties such as myelination and fibre density (Le Bihan and Johansen-Berg, 2012) and has clinical relevance in patient cohorts (Assaf and Pasternak, 2008; Dennis and Thompson, 2013*a*; Assaf *et al*., 2019). We used tract-based spatial statistics (TBSS) (Smith et al., 2006) to perform voxel-wise comparison of FA across the brain’s white matter, whilst controlling for multiple comparisons. We further investigated white matter connectivity by constructing structural brain networks, or connectomes (Sporns et al., 2005), in which nodes represent brain regions and edges were determined by probabilistic tractography. We characterised structural networks by drawing on techniques from graph theory (Bullmore and Sporns, 2009; Hagmann *et al*., 2010*a*; Fornito *et al*., 2013; Bassett and Sporns, 2017), allowing comparison of quantitative differences in whole-brain network structure. Such techniques have previously been used to characterise the developing human connectome (Hagmann *et al*., 2010*b*; Dennis and Thompson, 2013*b*; Morgan *et al*., 2018), as well as in the study of specific neurodevelopmental complications such as CP (Arrigoni et al., 2016; Pannek et al., 2014) and neurodevelopmental impairments following preterm birth (Brown et al., 2014; Muñoz-Moreno et al., 2016). We then used the network-based statistic (NBS) (Zalesky et al., 2012, 2010) to look for subsets of connections (subnetworks) which were weakened in cases, and subnetwork which related to measures of cognition.

Our findings demonstrate that, although TH reduces severe disabilities after NE, underlying structural deficits are present which are associated with the cognitive differences found between cases and controls at school-age. These differences are often overlooked as most children given TH for NE do not demonstrate significant deficits in performance at 18 months.

## 2 Materials and Methods

### 2.1 Participants

Informed and written consent was obtained from the parents of participants, in accordance with the Declaration of Helsinki. Ethical approval was obtained from the North Bristol Research Ethics Committee and the Health Research Authority (REC ID: 15/SW/0148).

#### 2.1.1 Cases

Eligibility criteria were as follows: gestation at birth ≥36 weeks and treatment with TH as standard clinical care based on TOBY trial eligibility criteria including signs of perinatal asphyxia and moderate to severe encephalopathy, confirmed by amplitude integrated electroencephalogram (Azzopardi et al., 2009). Children were excluded if they had started cooling later than six hours after birth, were cooled for less than three days, had received Xenon as part of a neuroprotective feasibility study, had been found to have a metabolic or genetic disorder, or if any major intracranial haemorrhage or structural brain abnormality could be seen on the neonatal MRI scan. Cases were sequentially selected from the cohort of children who received TH between 2008 and 2011. These data are maintained by the Bristol Neonatal Neurosciences group at St Michael’s Hospital, Bristol, UK, under previous ethics approval (REC ID: 09/H0106/3). A diagnosis of CP was ruled out at 2 years and reconfirmed at 6-8 years. Children were native English speakers and had no additional medical diagnosis other than NE.

#### 2.1.2 Controls

The control group consisted of children matched for age, sex and socio-economic status (Lee-Kelland et al., 2020). Children were excluded if they were born before 36 weeks gestation, had any history of NE or other medical issues of a neurological nature, or were not native English speakers.

### 2.2 Cognitive Assessment

Cognitive performance was assessed using the Wechsler Intelligence Scale for Children 4th Edition (WISC-IV) (Kaufman et al., 2006), which summarises raw score performance from 10 subsets into 10 scaled scores. These 10 scores are summed in four domains – verbal comprehension, perceptual reasoning, processing speed and working memory – which are combined to form a full-scale intelligence quotient (FSIQ) score. Cognitive testing was administered by assessors who were not previously involved with the patients’ care and were blinded to case-control status.

### 2.3 Image Acquisition

T1-weighted images and DWI data were acquired with a Siemens 3 tesla Magnetom Skyra MRI scanner at the Clinical Research and Imaging Centre (CRiCBristol), Bristol, UK. An experienced radiographer placed children supine within the 32-channel receive only head-coil, and head movement was minimised with memory-foam padding. Children wore earplugs and were able to watch a film of their choice. A volumetric T1-weighted anatomical scan was acquired for tissue segmentation and parcellation, with the magnetisation-prepared rapid acquisition gradient echo (MPRAGE) sequence using the following parameters: echo time (TE) = 2.19 ms; inversion time (TI) = 800 ms; repetition time (TR) = 1500 ms; flip angle = 9°; field of view (FoV) 234 × 250 mm; 240 slices; 1.0 mm isotropic voxels. DWI data were acquired for tractography and microstructural analysis, with a multiband echo-planar imaging (EPI) sequence, using the following parameters: TE = 70 ms; TR = 3150 ms; FoV 192 × 192 mm; 60 slices; 2.0 mm isotropic voxels, flip angle 90°, phase encoding in the anterior-posterior direction, in-plane acceleration factor = 2 (GRAPPA (Griswold et al., 2002)), through-plane multi-band factor = 2 (Moeller *et al*., 2010; Setsompop *et al*., 2012*a*, *b*). For the purpose of data averaging and eddy-current distortion correction, two sets of diffusion-weighted images were acquired with b = 1,000 s mm^-2^ in 60 diffusion directions, equally distributed according to an electrostatic repulsion model, as well as 8 interspersed b = 0 images, with one data set acquired with positive phase encoding steps, then repeated with negative steps (so-called, “blip-up, blip-down”), giving a total of 136 images.

### 2.4 Quality Control

The quality of the DWI data was assessed using the EddyQC tool (Bastiani et al., 2019) from the FMRIB Software Library (FSL, http://fsl.fmrib.ox.ac.uk) (Smith et al., 2004). Scans were rejected if the root-mean-square of all movement and eddy current metrics from EddyQC was greater than one standard deviation above the mean for all participants.

T1-weighted anatomical images were assessed visually; any scans with severe movement artefacts were rejected. The remaining scans were processed with the structural pipeline described below, followed by further visual inspection of the parcellation and tissue segmentation. Scans were further rejected at this stage if any moderate artefacts had caused errors in the parcellation or segmentation.

Figure 1 shows the process of recruitment and scan quality control. We recruited 51 cases and 43 controls for this study. Of these, 7 cases and 4 controls did not want to undergo scanning. A further 4 cases had incomplete data due to movement during the scan. DWI quality control led to the rejection of a further 6 cases and 2 controls. One further case and one control were rejected due to incorrect image volume placement. This left 33 case and 36 control scans which passed the DWI quality control, which were used in the TBSS analysis. Of these remaining 69 datasets, the anatomical scan for 11 cases and 4 controls was not of sufficient quality to allow segmentation and parcellation, leaving 22 cases and 32 controls for network analysis. Participant demographics are shown in Table 1.

**Table 1:** Participant demographics. Demographics are shown for each of the TBSS and network analysis groups. Median (range) is shown for age, socio-economic status and FSIQ. Socio-economic status is defined as follows: A = upper middle class, B = middle class, C1 = lower middle class, C2 = skilled working class, D = working class, E = casual worker or unemployed.

|  | TBSS |  |  | Network Analysis |  |  |
| --- | --- | --- | --- | --- | --- | --- |
|  | Cases<br>(n = 33) | Controls<br>(n = 36) | p | Cases<br>(n = 22) | Controls<br>(n = 32) | p |
| Age | 6.9 (6.0-7.9) | 7.0 (6.1-7.9) | 0.5555 | 7.0 (6.0-7.8) | 7.0 (6.1-7.8) | 0.5428 |
| M/F | 18/15 | 19/17 | 0.8894 | 12/10 | 16/16 | 0.7526 |
| Socio-economic status | C1 (A-E) | B (A-D) | 0.1568 | C1 (A-D) | B (A-D) | 0.0924 |
| FSIQ | 93 (62-115) | 108 (75-137) | <0.0001 | 98 (62-114) | 108 (75-137) | 0.0010 |
| Perceptual Reasoning | 90 (67-123) | 108 (84-145) | <0.0001 | 91 (67-110) | 108 (84-145) | <0.0001 |
| Processing Speed | 97 (68-136) | 106 (68-141) | 0.0787 | 98.5 (75-136) | 106 (68-141) | 0.1221 |
| Verbal Comprehension | 98 (73-126) | 109 (81-126) | 0.0012 | 98 (73-126) | 109 (81-126) | 0.0114 |
| Working Memory | 97 (62-116) | 105.5 (77-135) | 0.0076 | 94 (62-116) | 105.5 (77-135) | 0.0290 |

**Figure 1:**
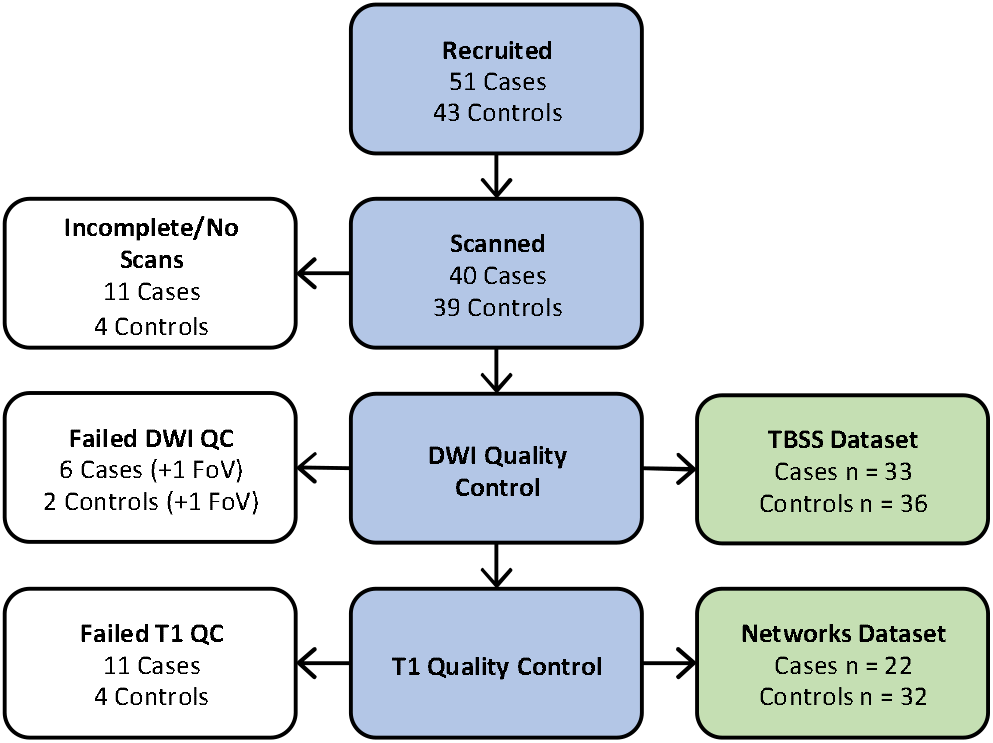
Recruitment. Flowchart of participants at each stage of quality control. FoV = field of view, indicating the scans which were rejected due to incorrect image volume placement.

Anatomical images were visually assessed for focal lesions and abnormal signal intensities. In the TBSS datasets, lesions were present in 1 case and 2 controls. In the network analysis datasets, lesions were present in 1 control. These lesions were judged by the blinded assessor (FC) to be non-severe, consequently these subjects were not excluded.

Note that previous findings from the same cohort demonstrate reduced performance in cases in all WISC-IV domains (Lee-Kelland et al., 2020), whereas in the smaller group which passed quality control in this study cases exhibit significantly reduced performance in perceptual reasoning, verbal comprehension, working memory and FSIQ. Though processing speed was reduced in cases in this study, the difference was not significant (see Table 1).

### 2.5 TBSS

Voxelwise statistical analysis of the FA data was carried out using TBSS (Smith et al., 2006), part of FSL. FA images were generated by fitting a tensor model to the diffusion data using FSL’s FDT software. All images were then nonlinearly registered to one subject, chosen automatically by finding the most representative subject, which was then affine registered to MNI152 standard space. This is the recommended procedure when testing data from children, which may not register well to an adult template (Smith et al., 2006). The mean FA image was then eroded to create a skeletonised representation of the white matter tracts. Each subject’s registered FA image was then projected onto this skeleton to allow voxelwise statistics.

### 2.6 Structural Network Construction

A weighted connectome was constructed for each subject, with nodes defined by parcellation of the anatomical scan and edges determined by probabilistic tractography using the DWI data. The processing pipeline, described in more detail below, is summarised in Figure 2.

**Figure 2:**
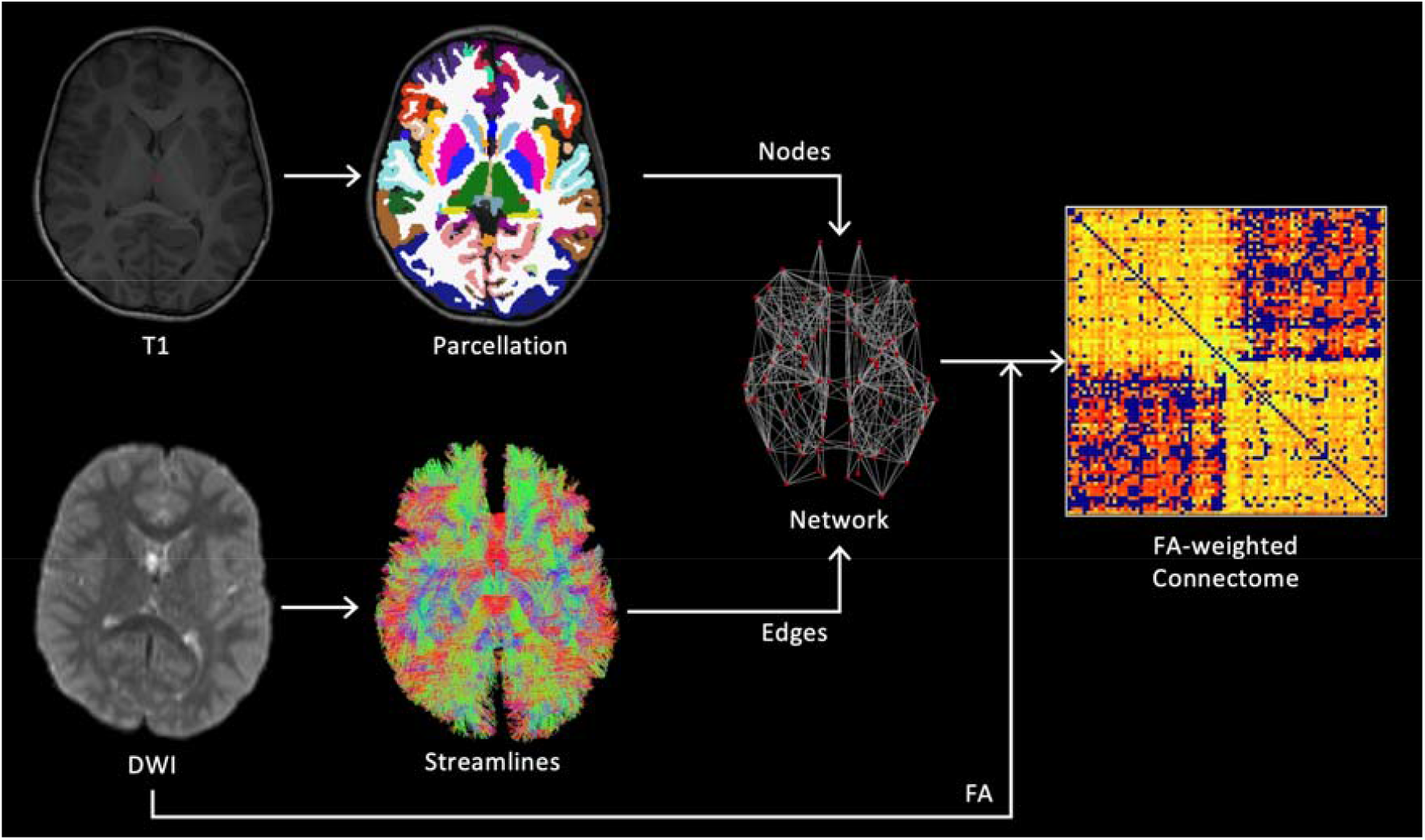
Processing pipeline. Method for constructing structural brain networks from T1 and DWI data. Cortical and sub-cortical nodes were defined by segmentation of the T1-weighted structural scan. Edges were determined by seeding streamlines from the cortical grey/white matter interface and performing tractography using the fibre orientation distribution obtained by spherical deconvolution of the measured diffusion signal. Edges were weighted by the mean FA along all streamlines passing between the corresponding pair of nodes, and the resulting network was represented by a connectivity matrix.

#### 2.6.1 Node Definition

T1-weighted anatomical images were denoised with the Advanced Normalization Tools DenoiseImage tool (http://github.com/ANTsX/ANTs) (Manjón et al., 2010). Brain extraction was performed with either SPM8-VBM (http://fil.ion.ucl.ac.uk/spm) (Ashburner and Friston, 2005) or CAT12 (http://www.neuro.uni-jena.de/cat) (Gaser and Dahnke, 2016) depending on which gave better delineation of the brain surface for each subject. Each subject’s T1-weighted image was parcellated, using FreeSurfer (http://surfer.nmr.mgh.harvard.edu) (Fischl, 2012), according to the Desikan-Killiany atlas (Desikan et al., 2006) (a total of 84 regions; 34 cortical, 7 subcortical and 1 cerebellar per hemisphere). The FIRST (Patenaude et al., 2011) subcortical segmentation tool from FSL was found to give better segmentation of subcortical structures (including the hippocampus and amygdala) than FreeSurfer, so this was combined with the cortical parcellation from FreeSurfer using the labelsgmfix tool from MRtrix (www.mrtrix.org) (Tournier et al., 2019).

#### 2.6.2 Edge Definition

DWI data were corrected for eddy current induced distortions and subject movements using EDDY (Andersson and Sotiropoulos, 2016) and TOPUP (Andersson et al., 2003), from FSL. Subsequent DWI processing and tractography steps were performed using MRtrix. The response function (the DWI signal for a typical fibre population) was estimated from the data (Tournier et al., 2013) in order to calculate the fibre orientation distribution (FOD) by performing constrained-spherical deconvolution of the response function from the measured DWI signal (Tournier et al., 2007). The normalised FOD image and the five-tissue-type segmentation of the T1-weighted anatomical image were used to perform anatomically-constrained tractography (Smith et al., 2012) using second-order integration over FODs (Tournier et al., 2010), with the following parameters: step size = 1 mm, minimum length = 50 mm, cutoff FOD magnitude = 0.1, maximum angle between steps = 30°. Streamlines were seeded in the interface between grey and white matter and only accepted if they terminated in subcortical or cortical grey matter. Terminated streamlines which were not accepted were allowed to backtrack to a valid point to be resampled (Smith et al., 2012). This method was used to generate 10 million streamlines which were subsequently filtered to 1 million using spherical-convolution informed filtering of tractograms (Smith et al., 2013) in order to improve biological plausibility and remove length bias. FA images were then used to assign a weight to each streamline according to the mean FA along its path. In order to construct a weighted graph for each subject, edges were defined between any pair of nodes connected by at least one streamline, with the connection strength defined by the mean FA along all streamlines connecting the nodes.

### 2.7 Network Metrics

We selected the following metrics to quantify properties of the FA-weighted structural connectivity networks: average strength, characteristic path length, global efficiency, local efficiency, clustering coefficient, modularity and small-worldness. These are defined below (for an in-depth description see Rubinov and Sporns, 2010).

The strength of a node is defined as the sum of the weights of all edges connected to the node. The average weight for the entire graph is equal to the average node strength across all nodes. The characteristic path length of the graph is the average of the shortest path from each node to every other node, where the edge distances used to calculate path lengths are defined inversely to edge weights (making stronger connections equivalent to shorter paths). Note that this does not reflect physical distance between regions in the brain. A shorter characteristic path length indicates stronger connectivity across brain regions, thus implying stronger potential for integration (Rubinov and Sporns, 2010). Global efficiency is the average of the inverse of the shortest path length. This has a roughly inverse relationship with characteristic path length, and therefore indicates integration (Bullmore and Sporns, 2009). However, the two metrics differ in the edges they are influenced by; the calculation of characteristic path length is more dependent on longer paths, whereas global efficiency is more dependent on shorter paths.

Local efficiency of a given node is the average of the inverse of the shortest path length between the immediate neighbours of that node. This is then averaged across all nodes to give a single measure for the whole graph. The clustering coefficient gives the number of connections between the nearest neighbours of a node as a fraction of the maximum number of possible connections. Modularity indicates how well the network can be split up into relatively separate communities (i.e. modules) of nodes by measuring a normalised ratio of the number of within-module connections to the number of between-module connections. Local efficiency, clustering coefficient and modularity indicate the efficiency of local information transfer, thus indicating the potential for segregated functional processing (Bullmore and Sporns, 2009; Rubinov and Sporns, 2010).

Both integration and segregation are required for brain networks to carry out localised and distributed processing simultaneously (Tononi et al., 1994). The degree to which a network exhibits both segregation and integration is measured by the small-worldness of the network (Muldoon et al., 2016; Rubinov and Sporns, 2010). A high degree of small-worldness is characterised by a high clustering coefficient and low characteristic path length compared to random graphs. We measured small-worldness with small-world propensity (Muldoon et al., 2016). All other metrics were calculated with the Brain Connectivity Toolbox (http://www.brain-connectivity-toolbox.net) (Rubinov and Sporns, 2010).

### 2.8 Statistical Analysis

Group differences between case and control network metrics were tested using two-tailed, unpaired t-tests. Correlation of network metrics with cognitive score was then tested by calculating the partial Pearson correlation coefficient, including age and sex as covariates. In order to reduce the effect of multiple comparisons and increase statistical power, each network metric was tested for correlation with FSIQ, not with every WISC-IV domain. Bonferroni correction was applied to correct for multiple comparisons. Statistical analysis of the network metrics was performed in MATLAB (R2018b, Mathworks). For TBSS, significance was tested using FSL’s non-parametric permutation testing software, RANDOMISE (Winkler et al., 2014). We used 10,000 permutations and applied threshold-free cluster enhancement to correct for multiple comparisons. Significant results have corrected p < 0.05.

#### 2.8.1 Network-Based Statistic (NBS)

We used NBS to test the hypothesis that cases exhibit reduced connectivity (i.e. reduced FA) compared to controls, based on previously reported findings of reduced FA in white matter in neonates treated with TH for NE (Lally et al., 2019; Tusor et al., 2012). We also explored group differences in the relationship between cognitive scores and connectivity.

NBS (Zalesky et al., 2012, 2010) is a nonparametric, permutation-based approach for controlling family-wise error rate (FWER) on the level of subnetworks. NBS identifies connected subnetworks in which each edge satisfies the given contrast (e.g. group differences in connectivity). The t-statistic is calculated for each edge in the network, then thresholded at a chosen value. Of the remaining suprathreshold edges, the size of each connected subnetwork (given by the number of edges) is stored. This process is repeated for random permutations of the data to estimate the null distribution. The FWER-corrected p-value for each subnetwork is given by the number of permutations for which the largest connected subnetwork in the permuted data is the same size or larger than the given subnetwork, normalised by the number of permutations.

We tested for reduced connectivity in cases compared to controls (one-tailed) and for group differences in the dependence of cognitive scores on edge weights (two-tailed). We tested all four cognitive domains for correlation (perceptual reasoning, processing speed, verbal comprehension, working memory) in addition to FSIQ. We used 10,000 permutations to calculate the p-value. In order to only test robust edges, only connections present in >50% of cases and >50% of controls were assessed. Age and sex were included as covariates in a general linear model in all tests (design matrices are shown in Supplementary Tables 2 and 3). As recommended in the literature (Zalesky et al., 2012, 2010), a range of t-statistic thresholds were tested (2.5-3.5) to find the value which gave robust results (Supplementary Figure 2). This procedure allows identification of large subnetworks with subtle effects (at low primary thresholds) as well as smaller subnetworks with strong effects (at high primary thresholds). Significant results have p < 0.05 (FWER-corrected).

### 2.9 Visualisation

Subnetworks were visualised with the BrainNet Viewer (https://www.nitrc.org/projects/bnv/) (Xia et al., 2013) and as Circos connectograms (http://www.circos.ca) (Krzywinski et al., 2009).

### 2.10 Data Availability

The data that support the findings in this article are available upon reasonable request to the corresponding author.

## 3 Results

### 3.1 TBSS

Figure 3 shows the results of voxelwise comparison of FA using TBSS, demonstrating widespread reduction in FA in cases compared to controls. The effect is most prominent in the fornix, the corpus callosum, anterior and posterior limbs of the internal capsule bilaterally, and the cingulum bilaterally, but can also be seen in other distributed areas of white matter. These results demonstrate extensive alterations to white matter microstructure in cases. This analysis was repeated with age and sex included as covariates in a general linear model; the results were largely unchanged (see Supplementary Figure 1).

**Figure 3:**
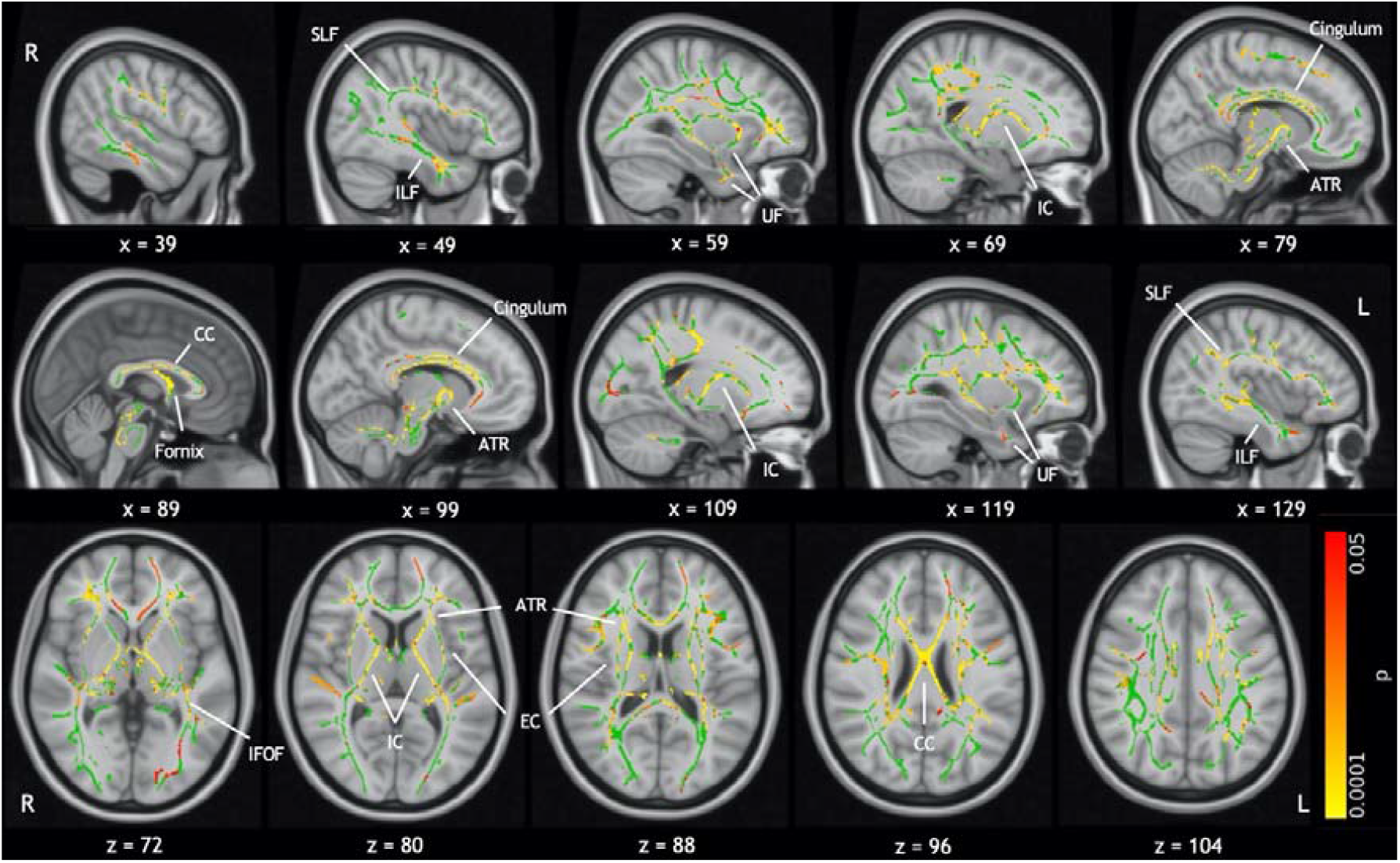
TBSS results. Results of voxelwise comparison of FA on the white matter skeleton (green). Significant results are indicated by the colour bar (p < 0.05, TFCE corrected). These are overlaid on the MNI standard template. Labels indicate some major white matter tracts and regions. Abbreviations are as follows: anterior thalamic radiation (ATR), corpus callosum (CC), external capsule (EC), internal capsule (IC), inferior fronto-occipital fasciculus (IFOF), inferior longitudinal fasciculus (ILF), superior longitudinal fasciculus (SLF), uncinate fasciculus (UF). Differences are most notable in the IC, CC, fornix and cingulum.

### 3.2 Network Metrics

#### 3.2.1 Group Differences

No significant group differences were found in network metrics (see Supplementary Table 1). Notably, small-world characteristics were expressed robustly across the entire cohort with all subjects expressing a small-world propensity greater than 0.82 (networks with small-world propensity > 0.6 are considered small-world (Muldoon et al., 2016)).

#### 3.2.2 Cognitive Correlations

Figure 4 shows the correlation of network metrics with FSIQ. In cases, FSIQ was significantly correlated with average node strength (r = 0.6858, p = 0.0059), local efficiency (r = 0.6320, p = 0.0196), global efficiency (r = 0.6672, p = 0.0092), clustering coefficient (r =0.6817, p = 0.0065) and characteristic path length (r = −0.6704, p = 0.0085). In controls, network metrics exhibited the same general trends as in cases, however none of the correlations were significant, despite there being a comparable spread in the residuals.

**Figure 4:**
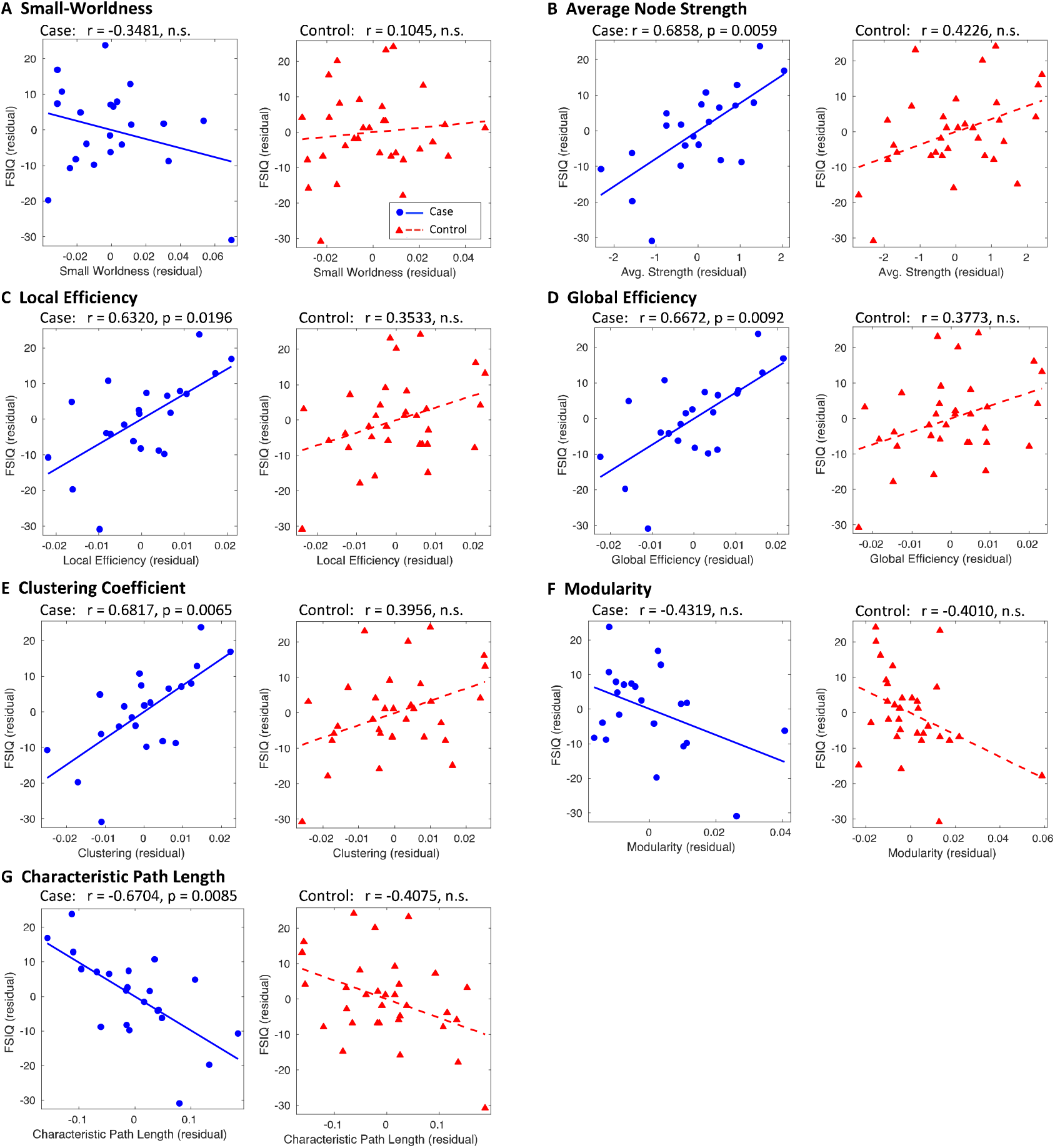
Correlation of network metrics with FSIQ. Network metrics and FSIQ were controlled for age and sex, with residuals plotted for cases (blue circles) and controls (red triangles). These are fitted with a blue solid line and red dashed line for cases and controls respectively. Where p > 0.05, plots are labelled as not significant (n.s.). p-values are Bonferroni corrected for the number of correlations performed.

#### 3.3 NBS

Figures 5 and 6 show the significant subnetworks identified by NBS. To reiterate; in each of the subnetworks, the tested contrast is expressed significantly at the level of each individual connection, with FWER controlled for the whole subnetwork. Significant results were found for: reduced connectivity (equating to reduced FA) in cases compared to controls; stronger relationship between connectivity and FSIQ in cases than in controls; and stronger relationship between connectivity and processing speed in cases than in controls. No results were found for group differences in the relationship between connectivity and perceptual reasoning, verbal comprehension or working memory.

**Figure 5:**
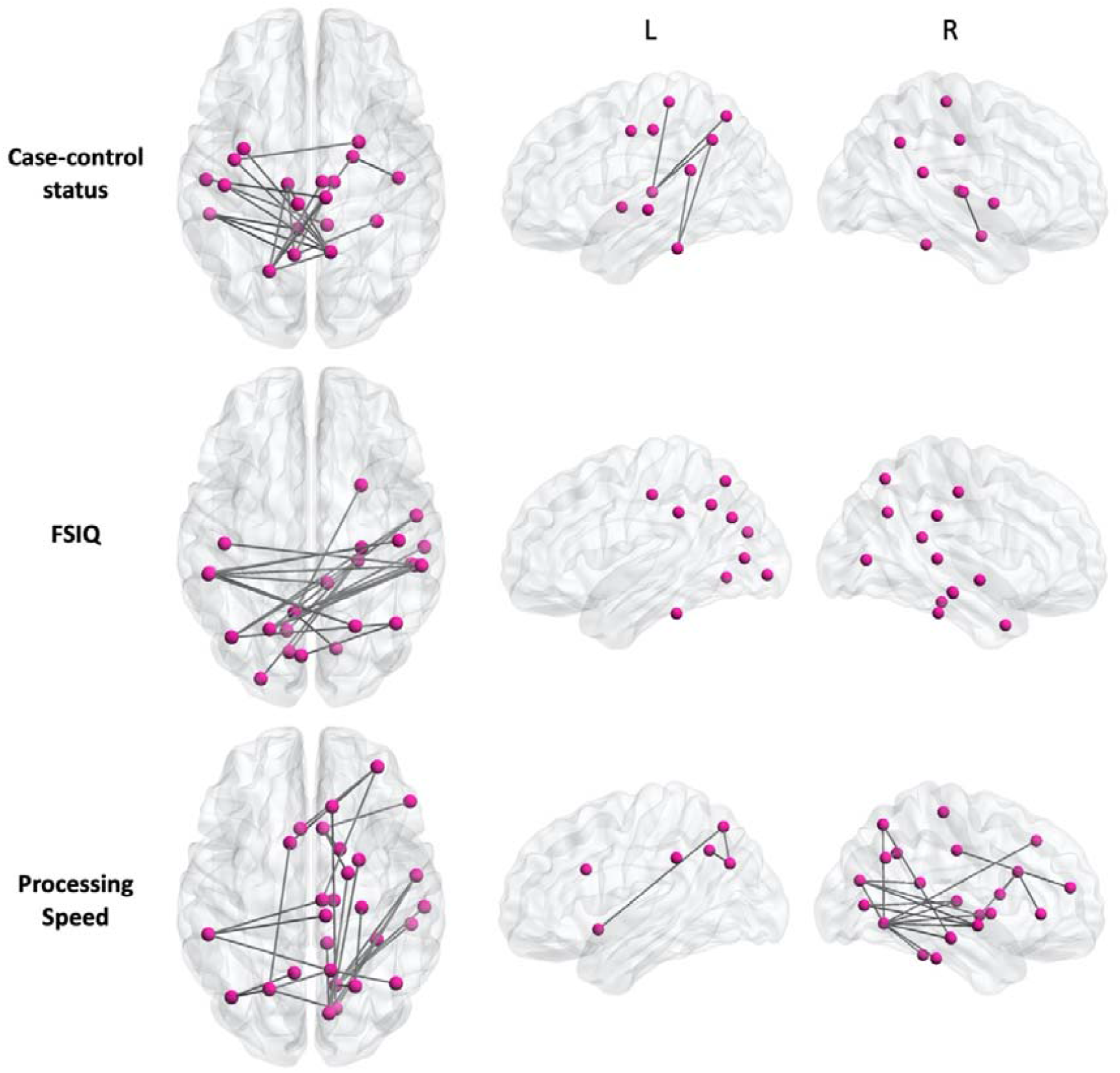
NBS results. Subnetworks are shown for case-control comparison (top), correlation with FSIQ (middle) and correlation with processing speed (bottom). In the case-control status subnetwork, all connections shown are significantly weakened in cases compared to controls (reflecting lower FA in cases). In the FSIQ and processing speed subnetworks, the dependence of cognitive score on connection strength is significantly higher in cases than in controls. The dorsal (axial) view shows all connections, while the lateral (sagittal) views of the left and right cortices show the intrahemispheric connections.

**Figure 6:**
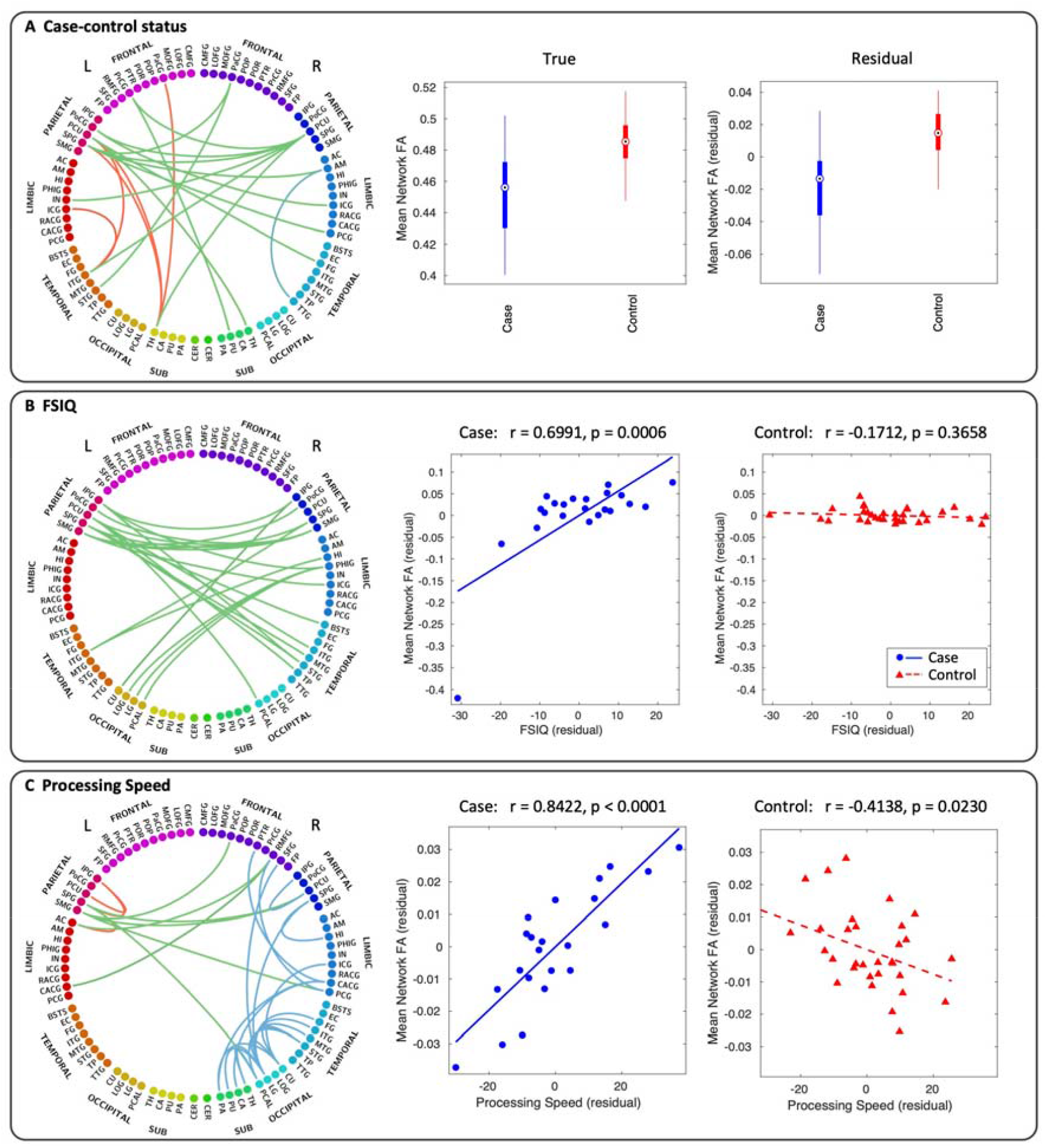
Subnetworks. Subnetworks given by NBS analysis of case-control comparison (A), correlation with FSIQ (B) and correlation with processing speed (C). Connectograms are shown with interhemispheric connections in green and intrahemispheric connections in red (left) and blue (right). Panel A also shows box plots of the mean FA across all connections in the case-control subnetwork for both the true FA values (left) and residual values after controlling for age and sex (right). In the box plots, the circle is the median, the solid box represents the 25^th^ to 75^th^ percentiles, and the lines extend to the minimum and maximum data points. Panels B and C also show scatter plots of the mean FA across all connections in the FSIQ and processing speed subnetwork, respectively, for both cases (blue circles, blue solid line) and controls (red triangles, red dashed line). The complete list of node label abbreviations is shown in the Supplementary Table 7.

Connectivity was significantly reduced in cases compared to controls (t = 2.8, p = 0.0304) in a subnetwork comprising 19 nodes (10 left, 9 right) and 20 edges (14 interhemispheric, 6 intrahemispheric). In this subnetwork, the five most well-connected nodes were the right precuneus cortex, left superior parietal gyrus, left precuneus cortex, left thalamus and left inferior temporal gyrus.

The relationship between connectivity and FSIQ was significantly stronger in cases than controls (t = 3.5, p = 0.0132) in a subnetwork comprising 23 nodes (10 left, 13 right) and 22 edges (all interhemispheric). The subnetwork was entirely composed of interhemispheric connections connecting parietal, limbic, temporal and occipital areas. The five most well-connected nodes were the left precuneus cortex, left and right supramarginal gyrus, left superior parietal gyrus and right parahippocampal gyrus.

The relationship between connectivity and processing speed was significantly stronger in cases than controls (t = 3.3, p = 0.0122) in a subnetwork comprising 28 nodes (6 left, 22 right) and 30 edges (7 interhemispheric, 23 intrahemispheric). The three most well-connected nodes were the right lingual gyrus, left superior parietal gyrus and right cuneus cortex. See Supplementary Tables 4-6 for the complete list of nodes in each subnetwork.

To provide graphical demonstration of each effect, the average FA in each of these subnetworks was calculated for each subject and plotted as a box plot for case-control differences (Fig. 6A) and plotted against FSIQ (Fig. 6B) and processing speed (Fig. 6C). This figure clearly demonstrates the effect captured by each subnetwork. The median of the subnetwork-averaged FA in cases was 6% lower in cases than in controls (p < 0.0001) in the case-control status subnetwork. The dependence of FSIQ on connectivity was much stronger in cases than controls in the FSIQ subnetwork (ANCOVA with age and sex as covariates; p < 0.0001). Similarly, the dependence of processing speed on connectivity was stronger in cases than controls in the processing speed subnetwork (ANCOVA with age and sex as covariates; p < 0.0001).

Removing the outlier in Figure 6B strengthened the correlation (r = 0.7860, p < 0.0001).

## Discussion

This study assessed white matter microstructure and connectivity properties in children aged 6-8 years who underwent TH for NE at birth and who did not develop CP, compared to a matched group of control children with no history of neurological issues. TBSS was used to compare white matter microstructural properties, derived from diffusion weighted imaging data at the voxel level, between cases and controls. Network analysis was used to further investigate the relationship between brain connectivity and cognitive measures in cases and controls, using graph theory to interpret connectome data. NBS was used to determine the specific connections associated with case-control status and those associated with cognitive performance.

Children who were treated with TH for NE at birth exhibited widespread reduction in FA compared with controls. Correlations with FSIQ were found in strength, local efficiency, global efficiency, clustering coefficient and characteristic path length of the structural networks, in cases only. NBS revealed subnetworks associated with case-control status, FSIQ and processing speed.

### 4.1 Cases Exhibit Widespread Alterations to White Matter Microstructure

Several factors can cause a reduction in FA, including reduced fibre density, cross-sectional area or myelination. Previous studies of neonates treated with TH for NE have investigated the relationship between white matter diffusion properties, measured in the first weeks following birth, and neurodevelopmental outcome at 2 years of age; these studies found a significant reduction in FA in infants with adverse outcomes, compared to those with favourable outcomes, in widespread areas of white matter including the centrum semiovale, corpus callosum, anterior and posterior limbs of the internal capsule, external capsules, fornix, cingulum, cerebral peduncles, optic radiations and inferior longitudinal fasciculus (Lally et al., 2019; Tusor et al., 2012). In addition, FA in many of these regions was found to correlate with developmental scores of children with and without CP (Tusor et al., 2012). Our findings in a select group of school-age children who did not develop CP, who had developmental scores in the normal range at 18 months and who were attending mainstream school, demonstrated reduced FA in many of the same areas of white matter as those highlighted in neonates, providing evidence that these microstructural differences persist to an older age group, even in the absence of CP. This suggests that children cooled for NE have an altered neurodevelopmental trajectory.

The question remains whether these alterations are caused by the cooling treatment, or if there is residual damage from the initial injury resulting from NE. There is conflicting evidence regarding the impact of TH on subcortical white matter. While one experimental study reported no adverse effect of hypothermia on subcortical white matter, brain maturation or neuronal death markers (Gressens et al., 2008), other studies have suggested that TH causes cell death in subcortical white matter (O’Brien et al., 2019; Wang et al., 2016). However, the damage resulting from NE without TH (Gao et al., 2012; Gosar et al., 2020; Ly et al., 2015; van Kooij et al., 2010, 2008), and the reduction in white matter lesions with TH compared to standard care following NE (Cheong et al., 2012; Rutherford et al., 2010) suggest that these microstructural alterations are likely attributable to the hypoxic-ischemic insult that preceded NE.

### 4.2 Structural Connectivity Correlates with Cognitive Outcome in Cases Only

We found no significant group mean differences in the network metrics. However, when considering cognitive performance, a close relationship was revealed between structural connectivity and functional outcome in cases only. In controls, though each metric exhibited the same general trend as in cases, none of the correlations with cognitive performance were significant, indicating that individual differences in structural connectivity play a bigger role in determining FSIQ in cases than in controls. The fact that this trend emerged in relation to cognitive performance, despite finding no significant group differences in network metrics, suggests that cases exhibit a broad spectrum of connectivity impairments, ranging from mild to severe, which relate to functional outcome in these children.

In cases, the positive correlation of global efficiency with FSIQ and negative correlation of characteristic path length with FSIQ indicate a relationship between cognitive performance and network integration, which reflects the brain’s ability to carry out distributed processing (Bullmore and Sporns, 2009; Rubinov and Sporns, 2010). Also in cases, the positive correlation of local efficiency and clustering coefficient with FSIQ demonstrate a relationship between cognitive performance and network segregation, which reflects localised processing capabilities (Rubinov and Sporns, 2010). These relationships provide further evidence for the link between the severity of connectivity impairment and cognitive outcome following the brain injury.

During development, increasing network segregation is thought to be associated with pruning, while increasing strength and integration are thought to be associated with myelination (Dennis and Thompson, 2013b; Tymofiyeva et al., 2014). We found an association between reduced cognitive performance and measures of segregation and integration, reinforcing the hypothesis that the developmental trajectory of the TH children is altered, potentially impacting the processes of myelination and pruning and resulting in a ceiling effect on functional outcome at school age.

Despite the association between FSIQ and network strength, efficiency, clustering and characteristic path length, no relationship was found with small-worldness or modularity. This suggests that brain reorganisation during development prioritises small-world, modular characteristics, such that no relationship emerges between these properties and the level of cognitive impairment resulting from NE. Similar findings have been reported in school-age children born extremely preterm or with intrauterine growth restriction (Fischi-Gomez et al., 2016).

### 4.3 Regions Involved in Attention and Visuo-spatial Processing have Impaired Connectivity in Cases

Connectivity, measured by FA, was significantly reduced in cases compared to controls in a subnetwork comprising several sensorimotor areas including the thalamus, putamen, precentral gyrus, postcentral gyrus, paracentral gyrus and the superior parietal gyrus. The superior parietal gyrus is concerned with aspects of attention and visuo-spatial perception, including the representation and manipulation of objects. The precuneus, which appears bilaterally as two of the three most well-connected nodes in the subnetwork, is associated with numerous highly integrated tasks including visuo-spatial imagery and episodic memory retrieval (Cavanna, 2007; Cavanna and Trimble, 2006). Other nodes in the subnetwork include the insula (sensorimotor as well as higher-level cognitive function (Uddin et al., 2017)), isthmus of the cingulate cortex (which has a role in memory), inferior temporal gyrus (visual processing and visual object recognition), superior temporal gyrus (visual information integration (Karnath, 2001; Shen et al., 2017)), fusiform gyrus (object and face recognition (Kleinhans et al., 2008; Pelphrey et al., 2007)), amygdala (emotional behaviour) and the posterior cingulate cortex (internally directed thought (Leech et al., 2011) and task management (Pearson et al., 2011)). The posterior cingulate cortex is also involved in controlling attention via interaction with the cognitive control network and has been linked to attentional impairments in brain injury, autism, attention deficit hyperactivity disorder and schizophrenia (Leech et al., 2011; Leech and Sharp, 2014). Both the precuneus and the posterior cingulate cortex feature in the default mode network (Raichle et al., 2001), suggesting a role in the neural correlates of consciousness (Cavanna, 2007).

The reduced connectivity to numerous regions involved in visuo-spatial processing and attention aligns with behavioural findings from a study by Tonks *et al*., demonstrating reduced visuo-spatial processing, attention difficulties and slower reaction times in this group of children (Tonks et al., 2019). Similarly, the sensorimotor regions included in the network (in particular the numerous thalamocortical connections) may account for the reduced motor performance in the absence of CP (Jary et al., 2019; Lee-Kelland et al., 2020) while the impaired connectivity to the amygdala may be linked to the increased likelihood of emotional behavioural difficulties (Lee-Kelland et al., 2020).

### 4.4 Connectivity to Regions Involved in Visuo-spatial Processing Correlates with Cognitive Outcome

Subnetworks were found in which there is a stronger dependence of aspects of cognitive outcome (FSIQ and processing speed) on connectivity in cases than in controls. Processing speed aims to measure the mental speed and cognitive flexibility of the child; however, the score is also affected by other cognitive factors such as visuo-motor coordination, visual discrimination, attention, short-term visual memory and concentration. FSIQ is a measure of the overall cognitive ability of an individual based on performance on all WISC-IV subtests (Kaufman et al., 2006). There were no edges common to the two subnetworks, indicating that the correlation with FSIQ was not driven by correlation with processing speed.

The most well-connected nodes in the FSIQ subnetwork are involved in visuo-spatial processing, memory and attention, but there are also connections to several association cortices and visual processing areas. All connections in the FSIQ subnetwork are interhemispheric, suggesting involvement of the corpus callosum. The processing speed subnetwork consists of predominantly visual processing regions, as well as areas involved in visuo-spatial function and attention, and sensorimotor areas. Importantly, the relationship between connectivity and outcome is significantly stronger in cases than in controls, as demonstrated in Figures 6B and C. This provides an extension to the idea of ceiling effects being imposed on the cognitive processing abilities of cases, whereby the connections in the subnetwork restrict cognitive outcome in cases, whereas the cognitive processing abilities of controls are less dependent on the strength of these particular connections.

Though perceptual reasoning, verbal comprehension and working memory were reduced in cases (see Table 1), group differences in the dependence of these domains on connectivity was not found. This could be due to the dependence of these domains on connectivity being equal across subjects regardless of case-control status, or due to these domains being dependent on different connections in each subject rather than on any distinct subnetwork. This may also be dependent on how well each WISC-IV domain reflects fundamental cognitive processes versus higher-level thinking.

### 4.5 Major Hubs in the Human Connectome are Among Those Affected in Cases

Several studies have investigated structural brain network properties to determine key, densely connected hub nodes which constitute a structural core, or “rich club”, of the human connectome (Gong et al., 2009; Hagmann et al., 2008; van den Heuvel and Sporns, 2011). These hub nodes are thought to play a central role in information integration.

These studies consistently identified the precuneus cortex as a key node in the rich club, as well as highlighting the posterior cingulate cortex, superior parietal cortex, paracentral lobule, isthmus of the cingulate cortex, superior temporal cortex and thalamus. Additionally, sensorimotor areas were among those found to be hubs during the neonatal period (Fransson et al., 2011; van den Heuvel et al., 2015) and have been shown to be affected in dyskinetic cerebral palsy (Ballester-Plané et al., 2017), which can also result from hypoxia at birth. Many of these rich club nodes were implicated in the relationship between connectivity and case-control status, FSIQ and processing speed.

It has been suggested that, due to their topological centrality and high biological cost, rich club nodes are particularly vulnerable to a wide range of pathogenic factors (Crossley et al., 2014; van den Heuvel and Sporns, 2013). The high metabolic rates of the precuneus cortex (Cavanna and Trimble, 2006) and posterior cingulate cortex (Leech and Sharp, 2014) support this suggestion of vulnerability. Increased vulnerability may be a reason for these nodes being implicated in NE children; these nodes are affected the most by the lack of oxygen during birth therefore they sustain lasting developmental alterations.

### 4.6 Strengths and Limitations

To our knowledge, this is the first study to investigate whole-brain structural connectivity in school-age children treated with TH for NE, who did not develop CP. We used a robust methodology of high angular resolution DWI combined with an anatomically-constrained tractography method capable of resolving crossing fibres. Movement can be a common issue when scanning children, therefore we applied a robust quality control pipeline. The rejection of scans due to movement artefact, as well as the incomplete or unobtained scans, resulted in a relatively small sample size. However, there were no significant differences between the cognitive scores of the rejected subjects and those included in the analysis. In order to increase the robustness of the NBS results, connections were only included in the analysis if expressed in >50% of cases and >50% of controls.

## 5 Conclusions

We demonstrate structural connectivity deficits relating to white matter microstructure and network connectivity properties in school-age children treated with TH for NE, who did not develop CP, compared to typically developing controls. We provide evidence for a relationship between structural connectivity and cognitive outcome and further demonstrate specific brain regions and connections which are associated with case-control status and with cognitive outcome. This work demonstrates that, despite the successes of TH in reducing cases of severe disability and death, there are still aspects of brain structure which are impacted by the process of NE despite treatment with TH. Further study involving neonatal scans and longitudinal investigation of the developmental aspects of these impairments could guide follow-up care and inform future therapeutic intervention strategies.

## Acknowledgements

We thank the children and their families for participating, Ngoc Jade Thai for her assistance with MR sequences, Aileen Wilson for her radiographical expertise, and Charlotte Whitfield and Emily Broadbridge for their assistance with neuropsychological assessment..

## References

Andersson, J.L.R., Skare, S., Ashburner, J., 2003. How to correct susceptibility distortions in spin-echo echo-planar images: application to diffusion tensor imaging. Neuroimage 20, 870–888. https://doi.org/10.1016/S1053-8119(03)00336-7

Andersson, J.L.R., Sotiropoulos, S.N., 2016. An integrated approach to correction for off-resonance effects and subject movement in diffusion MR imaging. Neuroimage 125, 1063–1078. https://doi.org/10.1016/J.NEUROIMAGE.2015.10.019

Arrigoni, F., Peruzzo, D., Gagliardi, C., Maghini, C., Colombo, P., Iammarrone, F.S., Pierpaoli, C., Triulzi, F., Turconi, A.C., 2016. Whole-Brain DTI Assessment of White Matter Damage in Children with Bilateral Cerebral Palsy: Evidence of Involvement beyond the Primary Target of the Anoxic Insult. Am. J. Neuroradiol. 37, 1347–1353. https://doi.org/10.3174/ajnr.A4717

Ashburner, J., Friston, K.J., 2005. Unified segmentation. Neuroimage 26, 839–851. https://doi.org/10.1016/j.neuroimage.2005.02.018

Assaf, Y., Johansen C: Berg, H., Thiebaut de Schotten, M., 2019. The role of diffusion MRI in neuroscience. NMR Biomed. 32, e3762. https://doi.org/10.1002/nbm.3762

Assaf, Y., Pasternak, O., 2008. Diffusion Tensor Imaging (DTI)-based White Matter Mapping in Brain Research: A Review. J. Mol. Neurosci. 34, 51–61. https://doi.org/10.1007/s12031-007-0029-0

Azzopardi, D., Strohm, B., Marlow, N., Brocklehurst, P., Deierl, A., Eddama, O., Goodwin, J., Halliday, H.L., Juszczak, E., Kapellou, O., Levene, M., Linsell, L., Omar, O., Thoresen, M., Tusor, N., Whitelaw, A., Edwards, A.D., 2014. Effects of Hypothermia for Perinatal Asphyxia on Childhood Outcomes. N. Engl. J. Med. 371, 140–149. https://doi.org/10.1056/NEJMoa1315788

Azzopardi, D. V., Strohm, B., Edwards, A.D., Dyet, L., Halliday, H.L., Juszczak, E., Kapellou, O., Levene, M., Marlow, N., Porter, E., Thoresen, M., Whitelaw, A., Brocklehurst, P., 2009. Moderate Hypothermia to Treat Perinatal Asphyxial Encephalopathy. N. Engl. J. Med. 361, 1349–1358. https://doi.org/10.1056/NEJMoa0900854

Ballester-Plané, J., Schmidt, R., Laporta-Hoyos, O., Junqué, C., Vázquez, é., Delgado, I., Zubiaurre-Elorza, L., Macaya, A., Póo, P., Toro, E., de Reus, M.A., van den Heuvel, M.P., Pueyo, R., 2017. Whole-brain structural connectivity in dyskinetic cerebral palsy and its association with motor and cognitive function. Hum. Brain Mapp. 38, 4594–4612. https://doi.org/10.1002/hbm.23686

Bassett, D.S., Sporns, O., 2017. Network neuroscience. Nat. Neurosci. 20, 353–364. https://doi.org/10.1038/nn.4502

Bastiani, M., Andersson, J.L.R., Cordero-Grande, L., Murgasova, M., Hutter, J., Price, A.N., Makropoulos, A., Fitzgibbon, S.P., Hughes, E., Rueckert, D., Victor, S., Rutherford, M., Edwards, A.D., Smith, S.M., Tournier, J.-D., Hajnal, J. V., Jbabdi, S., Sotiropoulos, S.N., 2019. Automated processing pipeline for neonatal diffusion MRI in the developing Human Connectome Project. Neuroimage 185, 750–763. https://doi.org/10.1016/J.NEUROIMAGE.2018.05.064

Brown, C.J., Miller, S.P., Booth, B.G., Andrews, S., Chau, V., Poskitt, K.J., Hamarneh, G., 2014. Structural network analysis of brain development in young preterm neonates. Neuroimage 101, 667–680. https://doi.org/10.1016/j.neuroimage.2014.07.030

Bullmore, E., Sporns, O., 2009. Complex brain networks: graph theoretical analysis of structural and functional systems. Nat. Rev. Neurosci. 10, 186–98. https://doi.org/10.1038/nrn2575

Cavanna, A.E., 2007. The Precuneus and Consciousness. CNS Spectr. 12, 545–552. https://doi.org/10.1017/S1092852900021295

Cavanna, A.E., Trimble, M.R., 2006. The precuneus: a review of its functional anatomy and behavioural correlates. Brain 129, 564–583. https://doi.org/10.1093/brain/awl004

Chakkarapani, E., Dingley, J., Liu, X., Hoque, N., Aquilina, K., Porter, H., Thoresen, M., 2010. Xenon enhances hypothermic neuroprotection in asphyxiated newborn pigs. Ann. Neurol. 68, 330–341. https://doi.org/10.1002/ana.22016

Cheong, J.L.Y., Coleman, L., Hunt, R.W., Lee, K.J., Doyle, L.W., Inder, T.E., Jacobs, S.E., Infant Cooling Evaluation Collaboration, for the, 2012. Prognostic Utility of Magnetic Resonance Imaging in Neonatal Hypoxic-Ischemic Encephalopathy. Arch. Pediatr. Adolesc. Med. 166. https://doi.org/10.1001/archpediatrics.2012.284

Crossley, N.A., Mechelli, A., Scott, J., Carletti, F., Fox, P.T., Mcguire, P., Bullmore, E.T., 2014. The hubs of the human connectome are generally implicated in the anatomy of brain disorders. Brain 137, 2382–2395. https://doi.org/10.1093/brain/awu132

Dennis, E.L., Thompson, P.M., 2013a. Typical and atypical brain development: a review of neuroimaging studies. Dialogues Clin. Neurosci. 15, 359–84.

Dennis, E.L., Thompson, P.M., 2013b. Mapping connectivity in the developing brain. Int. J. Dev. Neurosci. 31, 525–542. https://doi.org/10.1016/j.ijdevneu.2013.05.007

Desikan, R.S., Ségonne, F., Fischl, B., Quinn, B.T., Dickerson, B.C., Blacker, D., Buckner, R.L., Dale, A.M., Maguire, R.P., Hyman, B.T., Albert, M.S., Killiany, R.J., 2006. An automated labeling system for subdividing the human cerebral cortex on MRI scans into gyral based regions of interest. Neuroimage 31, 968–980. https://doi.org/10.1016/j.neuroimage.2006.01.021

Fischi-Gomez, E., Muñoz-Moreno, E., Vasung, L., Griffa, A., Borradori-Tolsa, C., Monnier, M., Lazeyras, F., Thiran, J.P., Hüppi, P.S., 2016. Brain network characterization of high-risk preterm-born school-age children. NeuroImage Clin. 11, 195–209. https://doi.org/10.1016/j.nicl.2016.02.001

Fischl, B., 2012. FreeSurfer. Neuroimage 62, 774–781. https://doi.org/10.1016/j.neuroimage.2012.01.021

Fornito, A., Zalesky, A., Breakspear, M., 2013. Graph analysis of the human connectome: Promise, progress, and pitfalls. Neuroimage 80, 426–444. https://doi.org/10.1016/j.neuroimage.2013.04.087

Fransson, P., Åden, U., Blennow, M., Lagercrantz, H., 2011. The Functional Architecture of the Infant Brain as Revealed by Resting-State fMRI. Cereb. Cortex 21, 145–154. https://doi.org/10.1093/cercor/bhq071

Gale, C., Statnikov, Y., Jawad, S., Uthaya, S.N., Modi, N., 2018. Neonatal brain injuries in England: population-based incidence derived from routinely recorded clinical data held in the National Neonatal Research Database. Arch. Dis. Child. - Fetal Neonatal Ed. 103, F301–F306. https://doi.org/10.1136/archdischild-2017-313707

Gao, J., Li, X., Hou, X., Ding, A., Chan, K.C., Qinli Sun, Wu, E.X., Jian Yang, 2012. Tract-based spatial statistics (TBSS): Application to detecting white matter tract variation in mild hypoxic-ischemic neonates, in: 2012 Annual International Conference of the IEEE Engineering in Medicine and Biology Society. IEEE, pp. 432–435. https://doi.org/10.1109/EMBC.2012.6345960

Gaser, C., Dahnke, R., 2016. CAT-a computational anatomy toolbox for the analysis of structural MRI data. HBM 2016 336–348.

Gong, G., He, Y., Concha, L., Lebel, C., Gross, D.W., Evans, A.C., Beaulieu, C., 2009. Mapping anatomical connectivity patterns of human cerebral cortex using in vivo diffusion tensor imaging tractography. Cereb. Cortex 19, 524–536. https://doi.org/10.1093/cercor/bhn102

Gosar, D., Tretnjak, V., Bregant, T., Neubauer, D., Derganc, M., 2020. Reduced white-matter integrity and lower speed of information processing in adolescents with mild and moderate neonatal hypoxic-ischaemic encephalopathy. Eur. J. Paediatr. Neurol. 28, 205–213. https://doi.org/10.1016/j.ejpn.2020.06.003

Gressens, P., Dingley, J., Plaisant, F., Porter, H., Schwendimann, L., Verney, C., Tooley, J., Thoresen, M., 2008. Analysis of Neuronal, Glial, Endothelial, Axonal and Apoptotic Markers Following Moderate Therapeutic Hypothermia and Anesthesia in the Developing Piglet Brain. Brain Pathol. 18, 10–20. https://doi.org/10.1111/j.1750-3639.2007.00095.x

Griswold, M.A., Jakob, P.M., Heidemann, R.M., Nittka, M., Jellus, V., Wang, J., Kiefer, B., Haase, A., 2002. Generalized autocalibrating partially parallel acquisitions (GRAPPA). Magn. Reson. Med. 47, 1202–1210. https://doi.org/10.1002/mrm.10171

Hagmann, P., Cammoun, L., Gigandet, X., Gerhard, S., Ellen Grant, P., Wedeen, V., Meuli, R., Thiran, J.P., Honey, C.J., Sporns, O., 2010a. MR connectomics: Principles and challenges. J. Neurosci. Methods 194, 34–45. https://doi.org/10.1016/j.jneumeth.2010.01.014

Hagmann, P., Cammoun, L., Gigandet, X., Meuli, R., Honey, C.J., Van Wedeen, J., Sporns, O., 2008. Mapping the structural core of human cerebral cortex. PLoS Biol. 6, 1479–1493. https://doi.org/10.1371/journal.pbio.0060159

Hagmann, P., Sporns, O., Madan, N., Cammoun, L., Pienaar, R., Wedeen, V.J., Meuli, R., Thiran, J.-P., Grant, P.E., 2010b. White matter maturation reshapes structural connectivity in the late developing human brain. Proc. Natl. Acad. Sci. 107, 19067–19072. https://doi.org/10.1073/pnas.1009073107

Jacobs, S.E., Berg, M., Hunt, R., Tarnow-Mordi, W.O., Inder, T.E., Davis, P.G., 2013. Cooling for newborns with hypoxic ischaemic encephalopathy. Cochrane Database Syst. Rev. https://doi.org/10.1002/14651858.CD003311.pub3

Jary, S., Lee-Kelland, R., Tonks, J., Cowan, F.M., Thoresen, M., Chakkarapani, E., 2019. Motor performance and cognitive correlates in children cooled for neonatal encephalopathy without cerebral palsy at school age. Acta Paediatr. Int. J. Paediatr. 108, 1773–1780. https://doi.org/10.1111/apa.14780

Jary, S., Smit, E., Liu, X., Cowan, F.M., Thoresen, M., 2015. Less severe cerebral palsy outcomes in infants treated with therapeutic hypothermia. Acta Paediatr. 104, 1241–1247. https://doi.org/10.1111/apa.13146

Karnath, H.-O., 2001. New insights into the functions of the superior temporal cortex. Nat. Rev. Neurosci. 2, 568–576. https://doi.org/10.1038/35086057

Kaufman, A.S., Flanagan, D.P., Alfonso, V.C., Mascolo, J.T., 2006. Test Review: Wechsler Intelligence Scale for Children, Fourth Edition (WISC-IV). J. Psychoeduc. Assess. 24, 278–295. https://doi.org/10.1177/0734282906288389

Kleinhans, N.M., Richards, T., Sterling, L., Stegbauer, K.C., Mahurin, R., Johnson, L.C., Greenson, J., Dawson, G., Aylward, E., 2008. Abnormal functional connectivity in autism spectrum disorders during face processing. Brain 131, 1000–1012. https://doi.org/10.1093/brain/awm334

Krzywinski, M., Schein, J., Birol, I., Connors, J., Gascoyne, R., Horsman, D., Jones, S.J., Marra, M.A., 2009. Circos: An information aesthetic for comparative genomics. Genome Res. 19, 1639–1645. https://doi.org/10.1101/gr.092759.109

Kyng, K.J., Skajaa, T., Kerrn-Jespersen, S., Andreassen, C.S., Bennedsgaard, K., Henriksen, T.B., 2015. A Piglet Model of Neonatal Hypoxic-Ischemic Encephalopathy. J. Vis. Exp. 1–12. https://doi.org/10.3791/52454

Lally, P.J., Montaldo, P., Oliveira, V., Soe, A., Swamy, R., Bassett, P., Mendoza, J., Atreja, G., Kariholu, U., Pattnayak, S., Sashikumar, P., Harizaj, H., Mitchell, M., Ganesh, V., Harigopal, Sundeep, Dixon, J., English, P., Clarke, P., Muthukumar, P., Satodia, P., Wayte, S., Abernethy, L.J., Yajamanyam, K., Bainbridge, A., Price, D., Huertas, A., Sharp, D.J., Kalra, V., Chawla, S., Shankaran, S., Thayyil, S., Lally, P.J., Montaldo, P., Oliveira, V., Soe, A., Swamy, R., Bassett, P., Mendoza, J., Atreja, G., Kariholu, U., Pattnayak, S., Sashikumar, P., Harizaj, H., Mitchell, M., Ganesh, V., Harigopal, Sundeeep, Dixon, J., English, P., Clarke, P., Muthukumar, P., Satodia, P., Wayte, S., Abernethy, L.J., Yajamanyam, K., Bainbridge, A., Price, D., Huertas, A., Sharp, D.J., Kalra, V., Chawla, S., Shankaran, S., Thayyil, S., 2019. Magnetic resonance spectroscopy assessment of brain injury after moderate hypothermia in neonatal encephalopathy: a prospective multicentre cohort study. Lancet Neurol. 18, 35–45. https://doi.org/10.1016/S1474-4422(18)30325-9

Le Bihan, D., Johansen-Berg, H., 2012. Diffusion MRI at 25: Exploring brain tissue structure and function. Neuroimage 61, 324–341. https://doi.org/10.1016/j.neuroimage.2011.11.006

Lee-Kelland, R., Jary, S., Tonks, J., Cowan, F.M., Thoresen, M., Chakkarapani, E., 2020. School-age outcomes of children without cerebral palsy cooled for neonatal hypoxic– ischaemic encephalopathy in 2008–2010. Arch. Dis. Child. - Fetal Neonatal Ed. 105, 8–13. https://doi.org/10.1136/archdischild-2018-316509

Leech, R., Kamourieh, S., Beckmann, C.F., Sharp, D.J., 2011. Fractionating the Default Mode Network: Distinct Contributions of the Ventral and Dorsal Posterior Cingulate Cortex to Cognitive Control. J. Neurosci. 31, 3217–3224. https://doi.org/10.1523/JNEUROSCI.5626-10.2011

Leech, R., Sharp, D.J., 2014. The role of the posterior cingulate cortex in cognition and disease. Brain 137, 12–32. https://doi.org/10.1093/brain/awt162

Ly, M.T., Nanavati, T.U., Frum, C.A., Pergami, P., 2015. Comparing tract-based spatial statistics and manual region-of-Interest labeling as diffusion analysis methods to detect white matter abnormalities in infants with hypoxic-Ischemic encephalopathy. J. Magn. Reson. Imaging 42, 1689–1697. https://doi.org/10.1002/jmri.24930

Manjón, J. V., Coupé, P., Martí-Bonmatí, L., Collins, D.L., Robles, M., 2010. Adaptive non-local means denoising of MR images with spatially varying noise levels. J. Magn. Reson. Imaging 31, 192–203. https://doi.org/10.1002/jmri.22003

Marlow, N., 2005. Neuropsychological and educational problems at school age associated with neonatal encephalopathy. Arch. Dis. Child. - Fetal Neonatal Ed. 90, F380–F387. https://doi.org/10.1136/adc.2004.067520

Martinez-Biarge, M., Bregant, T., Wusthoff, C.J., Chew, A.T.M., Diez-Sebastian, J., Rutherford, M.A., Cowan, F.M., 2012. White matter and cortical injury in hypoxic-ischemic encephalopathy: Antecedent factors and 2-year outcome. J. Pediatr. 161, 799–807. https://doi.org/10.1016/j.jpeds.2012.04.054

Moeller, S., Yacoub, E., Olman, C.A., Auerbach, E., Strupp, J., Harel, N., Uğurbil, K., 2010. Multiband multislice GE-EPI at 7 tesla, with 16-fold acceleration using partial parallel imaging with application to high spatial and temporal whole-brain fMRI. Magn. Reson. Med. 63, 1144–1153. https://doi.org/10.1002/mrm.22361

Morgan, S.E., White, S.R., Bullmore, E.T., Vértes, P.E., 2018. A Network Neuroscience Approach to Typical and Atypical Brain Development. Biol. Psychiatry Cogn. Neurosci. Neuroimaging 3, 754–766. https://doi.org/10.1016/j.bpsc.2018.03.003

Muldoon, S.F., Bridgeford, E.W., Bassett, D.S., 2016. Small-world propensity and weighted brain networks. Sci. Rep. 6, 1–13. https://doi.org/10.1038/srep22057

Muñoz-Moreno, E., Fischi-Gomez, E., Batalle, D., Borradori-Tolsa, C., Eixarch, E., Thiran, J.P., Gratacós, E., Hüppi, P.S., 2016. Structural brain network reorganization and social cognition related to adverse perinatal condition from infancy to early adolescence. Front. Neurosci. 10, 1–15. https://doi.org/10.3389/fnins.2016.00560

National Institute for Clinical Excellence (NICE), 2010. Therapeutic hypothermia withintracorporeal temperature monitoring for hypoxic perinatal brain injury [WWW Document]. URL https://www.nice.org.uk/guidance/ipg347 (accessed 6.18.20).

O’Brien, C.E., Santos, P.T., Kulikowicz, E., Reyes, M., Koehler, R.C., Martin, L.J., Lee, J.K., 2019. Hypoxia-Ischemia and Hypothermia Independently and Interactively Affect Neuronal Pathology in Neonatal Piglets with Short-Term Recovery. Dev. Neurosci. 41, 17–33. https://doi.org/10.1159/000496602

O’Connor, C.M., Ryan, C.A., Boylan, G.B., Murray, D.M., 2017. The ability of early serial developmental assessment to predict outcome at 5 years following neonatal hypoxic-ischaemic encephalopathy. Early Hum. Dev. 110, 1–8. https://doi.org/10.1016/j.earlhumdev.2017.04.006

Pannek, K., Boyd, R.N., Fiori, S., Guzzetta, A., Rose, S.E., 2014. Assessment of the structural brain network reveals altered connectivity in children with unilateral cerebral palsy due to periventricular white matter lesions. NeuroImage Clin. 5, 84–92. https://doi.org/10.1016/j.nicl.2014.05.018

Patenaude, B., Smith, S.M., Kennedy, D.N., Jenkinson, M., 2011. A Bayesian model of shape and appearance for subcortical brain segmentation. Neuroimage 56, 907–922. https://doi.org/10.1016/j.neuroimage.2011.02.046

Pearson, J.M., Heilbronner, S.R., Barack, D.L., Hayden, B.Y., Platt, M.L., 2011. Posterior cingulate cortex: adapting behavior to a changing world. Trends Cogn. Sci. 15, 143–151. https://doi.org/10.1016/j.tics.2011.02.002

Pelphrey, K.A., Morris, J.P., McCarthy, G., LaBar, K.S., 2007. Perception of dynamic changes in facial affect and identity in autism. Soc. Cogn. Affect. Neurosci. 2, 140–149. https://doi.org/10.1093/scan/nsm010

Raichle, M.E., MacLeod, A.M., Snyder, A.Z., Powers, W.J., Gusnard, D.A., Shulman, G.L., 2001. A default mode of brain function. Proc. Natl. Acad. Sci. 98, 676–682. https://doi.org/10.1073/pnas.98.2.676

Robertson, C.M.T., Finer, N.N., Grace, M.G.A., 1989. School performance of survivors of neonatal encephalopathy associated with birth asphyxia at term. J. Pediatr. 114, 753–760. https://doi.org/10.1016/S0022-3476(89)80132-5

Rubinov, M., Sporns, O., 2010. Complex network measures of brain connectivity: uses and interpretations. Neuroimage 52, 1059–69. https://doi.org/10.1016/j.neuroimage.2009.10.003

Rutherford, M., Ramenghi, L.A., Edwards, A.D., Brocklehurst, P., Halliday, H., Levene, M., Strohm, B., Thoresen, M., Whitelaw, A., Azzopardi, D., 2010. Assessment of brain tissue injury after moderate hypothermia in neonates with hypoxic–ischaemic encephalopathy: a nested substudy of a randomised controlled trial. Lancet Neurol. 9, 39–45. https://doi.org/10.1016/S1474-4422(09)70295-9

Setsompop, K., Cohen-Adad, J., Gagoski, B.A., Raij, T., Yendiki, A., Keil, B., Wedeen, V.J., Wald, L.L., 2012a. Improving diffusion MRI using simultaneous multi-slice echo planar imaging. Neuroimage 63, 569–580. https://doi.org/10.1016/j.neuroimage.2012.06.033

Setsompop, K., Gagoski, B.A., Polimeni, J.R., Witzel, T., Wedeen, V.J., Wald, L.L., 2012b. Blipped-controlled aliasing in parallel imaging for simultaneous multislice echo planar imaging with reduced g-factor penalty. Magn. Reson. Med. 67, 1210–1224. https://doi.org/10.1002/mrm.23097

Shen, W., Yuan, Y., Liu, C., Luo, J., 2017. The roles of the temporal lobe in creative insight: an integrated review. Think. Reason. 23, 321–375. https://doi.org/10.1080/13546783.2017.1308885

Smith, R.E., Tournier, J.-D., Calamante, F., Connelly, A., 2013. SIFT: Spherical-deconvolution informed filtering of tractograms. Neuroimage 67, 298–312. https://doi.org/10.1016/j.neuroimage.2012.11.049

Smith, R.E., Tournier, J.-D., Calamante, F., Connelly, A., 2012. Anatomically-constrained tractography: Improved diffusion MRI streamlines tractography through effective use of anatomical information. Neuroimage 62, 1924–1938. https://doi.org/10.1016/j.neuroimage.2012.06.005

Smith, S.M., Jenkinson, M., Johansen-Berg, H., Rueckert, D., Nichols, T.E., Mackay, C.E., Watkins, K.E., Ciccarelli, O., Cader, M.Z., Matthews, P.M., Behrens, T.E.J., 2006. Tract-based spatial statistics: voxelwise analysis of multi-subject diffusion data. Neuroimage 31, 1487–505. https://doi.org/10.1016/j.neuroimage.2006.02.024

Smith, S.M., Jenkinson, M., Woolrich, M.W., Beckmann, C.F., Behrens, T.E.J., Johansen-Berg, H., Bannister, P.R., De Luca, M., Drobnjak, I., Flitney, D.E., Niazy, R.K., Saunders, J., Vickers, J., Zhang, Y., De Stefano, N., Brady, J.M., Matthews, P.M., 2004. Advances in functional and structural MR image analysis and implementation as FSL. Neuroimage 23, S208–S219. https://doi.org/10.1016/J.NEUROIMAGE.2004.07.051

Sporns, O., Tononi, G., Kötter, R., 2005. The human connectome: A structural description of the human brain. PLoS Comput. Biol. 1, 0245–0251. https://doi.org/10.1371/journal.pcbi.0010042

Tonks, J., Cloke, G., Lee-Kelland, R., Jary, S., Thoresen, M., Cowan, F.M., Chakkarapani, E., 2019. Attention and visuo-spatial function in children without cerebral palsy who were cooled for neonatal encephalopathy: a case-control study. Brain Inj. 33, 894–898. https://doi.org/10.1080/02699052.2019.1597163

Tononi, G., Sporns, O., Edelman, G.M., 1994. A measure for brain complexity: relating functional segregation and integration in the nervous system. Proc. Natl. Acad. Sci. U. S. A. 91, 5033–7.

Tournier, J.-D., Calamante, F., Connelly, A., 2013. Determination of the appropriate b value and number of gradient directions for high-angular-resolution diffusion-weighted imaging. NMR Biomed. 26, 1775–1786. https://doi.org/10.1002/nbm.3017

Tournier, J.-D., Calamante, F., Connelly, A., 2010. Improved probabilistic streamlines tractography by 2nd order integration over fibre orientation distributions, in: Proceedings of the International Society for Magnetic Resonance in Medicine. p. 1670.

Tournier, J.-D., Calamante, F., Connelly, A., 2007. Robust determination of the fibre orientation distribution in diffusion MRI: Non-negativity constrained super-resolved spherical deconvolution. Neuroimage 35, 1459–1472. https://doi.org/10.1016/j.neuroimage.2007.02.016

Tournier, J.-D., Smith, R., Raffelt, D., Tabbara, R., Dhollander, T., Pietsch, M., Christiaens, D., Jeurissen, B., Yeh, C.-H., Connelly, A., 2019. MRtrix3: A fast, flexible and open software framework for medical image processing and visualisation. Neuroimage 202, 116137. https://doi.org/10.1016/j.neuroimage.2019.116137

Tusor, N., Wusthoff, C., Smee, N., Merchant, N., Arichi, T., Allsop, J.M., Cowan, F.M., Azzopardi, D., Edwards, A.D., Counsell, S.J., 2012. Prediction of neurodevelopmental outcome after hypoxic–ischemic encephalopathy treated with hypothermia by diffusion tensor imaging analyzed using tract-based spatial statistics. Pediatr. Res. 72, 63–69. https://doi.org/10.1038/pr.2012.40

Tymofiyeva, O., Hess, C.P., Xu, D., Barkovich, A.J., 2014. Structural MRI connectome in development: Challenges of the changing brain. Br. J. Radiol. 87. https://doi.org/10.1259/bjr.20140086

Uddin, L.Q., Nomi, J.S., Hébert-Seropian, B., Ghaziri, J., Boucher, O., 2017. Structure and Function of the Human Insula. J. Clin. Neurophysiol. 34, 300–306. https://doi.org/10.1097/WNP.0000000000000377

van den Heuvel, M.P., Kersbergen, K.J., de Reus, M.A., Keunen, K., Kahn, R.S., Groenendaal, F., de Vries, L.S., Benders, M.J.N.L., 2015. The Neonatal Connectome During Preterm Brain Development. Cereb. Cortex 25, 3000–3013. https://doi.org/10.1093/cercor/bhu095

van den Heuvel, M.P., Sporns, O., 2013. Network hubs in the human brain. Trends Cogn. Sci. 17, 683–696. https://doi.org/10.1016/j.tics.2013.09.012

van den Heuvel, M.P., Sporns, O., 2011. Rich-Club Organization of the Human Connectome. J. Neurosci. 31, 15775–15786. https://doi.org/10.1523/JNEUROSCI.3539-11.2011

van Kooij, B.J.M., van Handel, M., Nievelstein, R.A.J., Groenendaal, F., Jongmans, M.J., de Vries, L.S., 2010. Serial MRI and Neurodevelopmental Outcome in 9- to 10-Year-Old Children with Neonatal Encephalopathy. J. Pediatr. 157, 221-227.e2. https://doi.org/10.1016/j.jpeds.2010.02.016

van Kooij, B.J.M., van Handel, M., Uiterwaal, C.S.P.M., Groenendaal, F., Nievelstein, R.A.J., Rademaker, K.J., Jongmans, M.J., de Vries, L.S., 2008. Corpus Callosum Size in Relation to Motor Performance in 9-to 10-Year-Old Children with Neonatal Encephalopathy. Pediatr. Res. 63, 103–108. https://doi.org/10.1203/PDR.0b013e31815b4435

Wang, B., Armstrong, J.S., Reyes, M., Kulikowicz, E., Lee, J.-H., Spicer, D., Bhalala, U., Yang, Z.-J., Koehler, R.C., Martin, L.J., Lee, J.K., 2016. White matter apoptosis is increased by delayed hypothermia and rewarming in a neonatal piglet model of hypoxic ischemic encephalopathy. Neuroscience 316, 296–310. https://doi.org/10.1016/j.neuroscience.2015.12.046

Winkler, A.M., Ridgway, G.R., Webster, M.A., Smith, S.M., Nichols, T.E., 2014. Permutation inference for the general linear model. Neuroimage 92, 381–397. https://doi.org/10.1016/j.neuroimage.2014.01.060

Xia, M., Wang, J., He, Y., 2013. BrainNet Viewer: A Network Visualization Tool for Human Brain Connectomics. PLoS One 8, e68910. https://doi.org/10.1371/journal.pone.0068910

Yue, X., Mehmet, H., Penrice, J., Cooper, C., Cady, E., Wyatt, J.S., Reynolds, E.O.R., Edwards, A.D., Squier, M. V., 1997. Apoptosis and necrosis in the newborn piglet brain following transient cerebral hypoxia-ischaemia. Neuropathol. Appl. Neurobiol. 23, 16–25. https://doi.org/10.1111/j.1365-2990.1997.tb01181.x

Zalesky, A., Cocchi, L., Fornito, A., Murray, M.M., Bullmore, E., 2012. Connectivity differences in brain networks. Neuroimage 60, 1055–1062. https://doi.org/10.1016/j.neuroimage.2012.01.068

Zalesky, A., Fornito, A., Bullmore, E.T., 2010. Network-based statistic: Identifying differences in brain networks. Neuroimage 53, 1197–1207. https://doi.org/10.1016/j.neuroimage.2010.06.041

