## Supplementary Materials for "Disrupted Brain Connectivity in Children Treated with Therapeutic Hypothermia for Neonatal Encephalopathy"

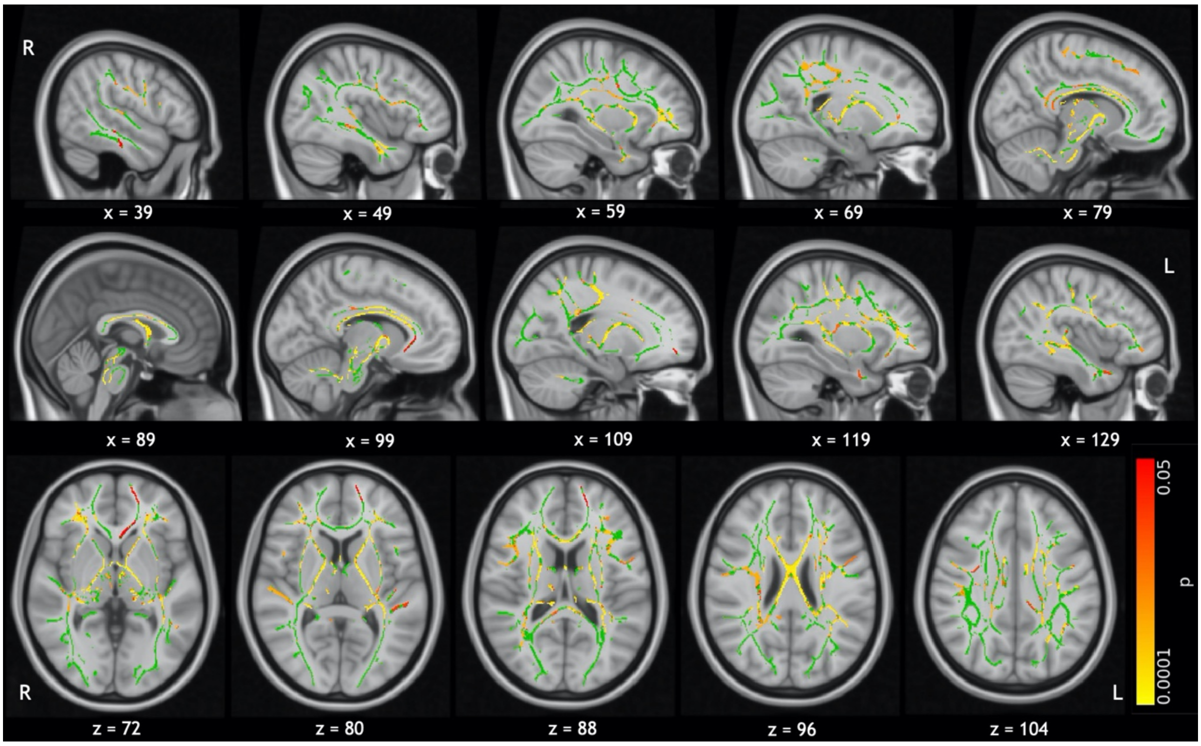


**Supplementary Figure 1: TBSS results with covariates.** FA on the white matter skeleton (green) was compared, with age and sex included as covariates in a general linear model. Significant results are indicated by the colour bar ($p < 0.05$, TFCE corrected). These are overlaid on the MNI standard template.

| Network Metric | Case | Control | Uncorrected p |
|---|---|---|---|
| Small-Worldness | 0.8594 | 0.8506 | 0.2347 |
| Average Node Strength | 28.07 | 28.74 | 0.0769 |
| Local Efficiency | 0.3925 | 0.3993 | 0.1072 |
| Global Efficiency | 0.3885 | 0.3954 | 0.0927 |
| Clustering Coefficient | 0.3572 | 0.3652 | 0.0684 |
| Modularity | 0.0682 | 0.0655 | 0.3286 |
| Characteristic Path Length | 2.740 | 2.689 | 0.0829 |

**Supplementary Table 1: Case-control comparison of network metrics.** Network metrics were calculated from the FA-weighted structural network of each subject. All network metrics were found to be normally distributed within each group (Kolmogorov-Smirnov test; $p < 0.05$). Columns show average network metrics for each group and the p-value calculated by a two-tailed, unpaired t-test. None of these differences were significant before correction so Bonferroni corrected p-values are not shown.

| Case | Control | Age | Sex |
|---|---|---|---|
| ***Design*** | | | |
| 1 | 0 | 6.75 | 0 |
| 0 | 1 | 7.25 | 1 |
| … | | | |
| ***Contrast*** | | | |
| -1 | 1 | 0 | 0 |

**Supplementary Table 2: Case-control comparison design matrix.** The design matrix and contrast to test for reduced connectivity in cases compared to controls, with age and sex included as covariates, are shown.

| Case | Control | Metric (case) | Metric (control) | Age | Sex |
|---|---|---|---|---|---|
| ***Design*** | | | | | |
| 1 | 0 | -7.5 | 0 | 6.75 | 0 |
| 0 | 1 | 0 | 5.47 | 7.25 | 1 |
| … | | | | | |
| ***Contrasts*** | | | | | |
| 0 | 0 | 1 | -1 | 0 | 0 |
| 0 | 0 | -1 | 1 | 0 | 0 |

**Supplementary Table 3: Correlation design matrix.** The design matrix and contrasts to test for stronger dependence of a given metric (e.g. FSIQ) on connectivity in cases than in controls, or a stronger dependence in controls than in cases, are shown. The metric is demeaned within each group and age and sex are included as covariates.

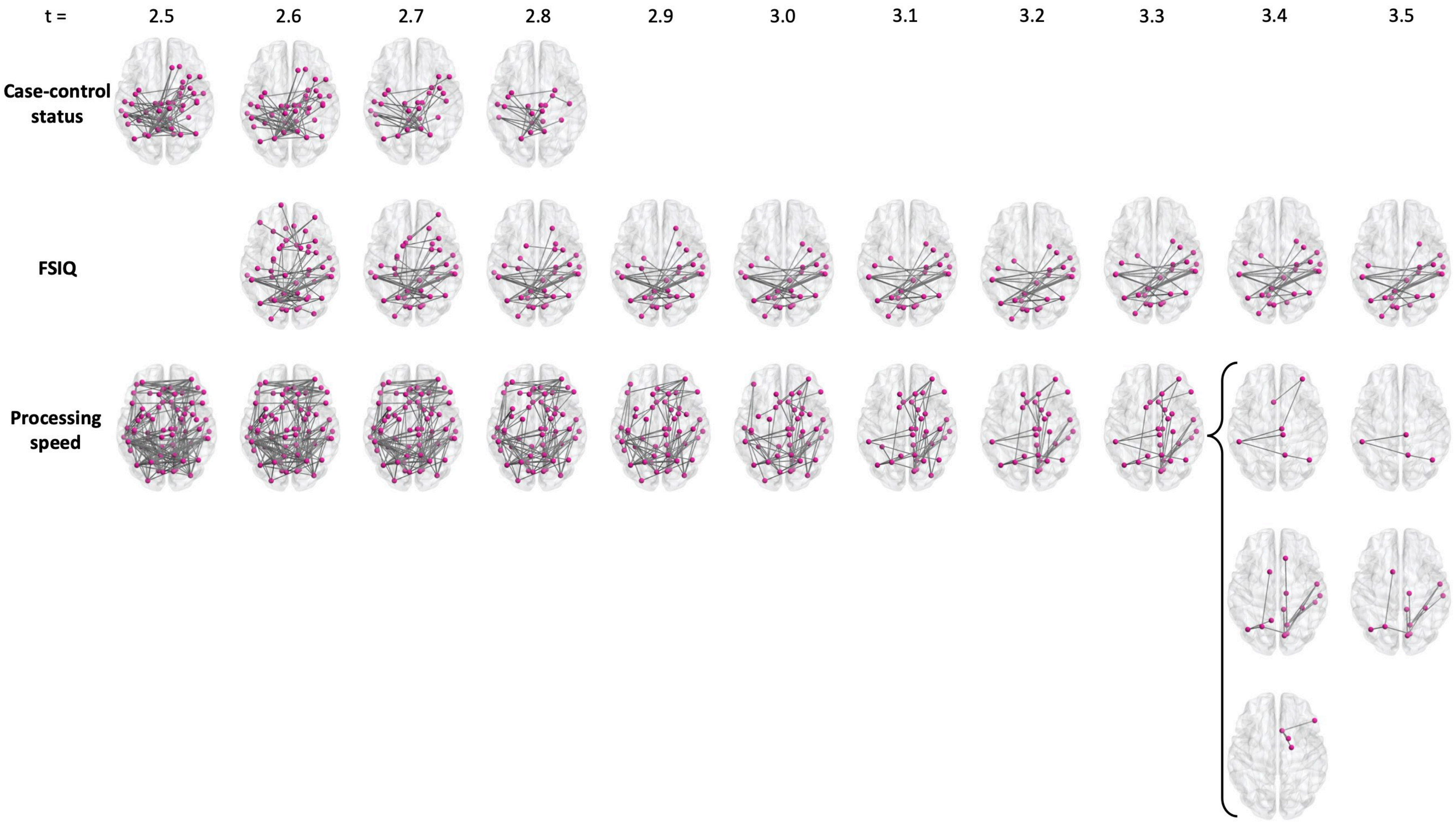


**Supplementary Figure 2: Testing t-statistic thresholds for NBS.** All networks shown are significant ($p < 0.05$). The primary thresholds chosen were: case-control status t = 2.8; FSIQ t = 3.5; processing speed t = 3.3.

| Region | Connections | Function |
|---|---|---|
| R precuneus* | 6 | Visuo-spatial imagery, episodic memory retrieval and "self-processing" operations, namely first-person perspective taking and an experience of agency (Cavanna, 2007; Cavanna and Trimble, 2006). |
| L superior parietal gyrus | 5 | Attention and visuo-spatial perception. |
| L precuneus* | 4 | See R precuneus above. |
| L thalamus* | 4 | Relaying sensorimotor signals to the cortex. |
| L inferior temporal gyrus | 3 | Visual processing and visual object recognition. |
| L postcentral gyrus | 2 | Primary somatosensory cortex. |
| L precentral gyrus | 2 | Primary motor area. |
| R amygdala | 2 | Emotional behaviour. |
| R paracentral gyrus* | 2 | Sensorimotor functions of the lower limb. |
| L isthmus of the cingulate gyrus* | 1 | Connects the posterior cingulate cortex to the hippocampus and has a role in memory. |
| L paracentral gyrus* | 1 | See R paracentral gyrus above. |
| L superior temporal gyrus | 1 | Visual information integration (Karnath, 2001; Shen et al., 2017). |
| L insula | 1 | Sensorimotor as well as higher-level cognitive function (Uddin et al., 2017). |
| R thalamus* | 1 | See L thalamus above. |
| R putamen | 1 | Movement regulation. |
| R fusiform gyrus | 1 | Object and face recognition (Kleinhans et al., 2008; Pelphrey et al., 2007). |
| R isthmus of the cingulate gyrus* | 1 | See L isthmus above. |
| R posterior cingulate gyrus | 1 | Internally directed thought (Leech et al., 2011) and task management (Pearson et al., 2011). |
| R transverse temporal gyrus | 1 | Part of the auditory cortex. |

**Supplementary Table 4: Nodes in the case-control status subnetwork.** These are listed by the number of connections they make within this subnetwork, and associated function. * nodes which are present bilaterally in the subnetwork.

| Region | Connections | Function |
|---|---|---|
| L precuneus cortex | 4 | Visuo-spatial imagery and episodic memory (see Supplementary Table 4). |
| L superior parietal gyrus* | 4 | Attention and visuo-spatial perception. |
| L supramarginal gyrus* | 4 | Visual word recognition and semantic word processing (Stoeckel et al., 2009). |
| R supramarginal gyrus* | 4 | See L supramarginal gyrus above. |
| R parahippocampal gyrus | 4 | Memory encoding and retrieval (Eichenbaum, 2000). |
| L inferior parietal gyrus* | 3 | Association area at the temporo-parietal junction (Igelström and Graziano, 2017). |
| R superior parietal gyrus* | 2 | See L superior parietal gyrus above. |
| R inferior parietal gyrus* | 2 | See L inferior parietal gyrus above. |
| L inferior temporal gyrus* | 2 | Visual processing and visual object recognition. |
| R superior temporal gyrus | 2 | Visual information integration (Karnath, 2001; Shen et al., 2017). |
| R inferior temporal gyrus* | 1 | See R inferior temporal gyrus above. |
| L cuneus cortex | 1 | Basic visual processing. |
| L lateral occipital gyrus | 1 | Visual object recognition (Grill-Spector et al., 1999). |
| L lingual gyrus | 1 | Visual association cortex. |
| L pericalcarine cortex* | 1 | Visual association cortex. |
| R pericalcarine cortex* | 1 | See L pericalcarine cortex above. |
| R hippocampus | 1 | Memory formation and spatial navigation (Eichenbaum, 2000). |
| R banks of superior temporal sulcus | 1 | Visual attention and goal-direction action (Shultz et al., 2011). |
| R isthmus of the cingulate gyrus | 1 | Connects the posterior cingulate cortex to the hippocampus and has a role in memory. |
| R middle temporal gyrus | 1 | Semantic memory processing. |
| R postcentral gyrus* | 1 | Primary somatosensory cortex. |
| L postcentral gyrus* | 1 | See R postcentral gyrus above. |
| R temporal pole | 1 | Social and emotional processing. |

**Supplementary Table 5: Nodes in the FSIQ subnetwork.** These are listed by the number of connections they make within this subnetwork, and associated function. * nodes which are present bilaterally in the subnetwork.

| Region | Connections | Function |
|---|---|---|
| R lingual gyrus | 9 | Visual association cortex. |
| L superior parietal gyrus* | 4 | Attention and visuo-spatial perception. |
| R cuneus cortex | 4 | Basic visual processing. |
| L supramarginal gyrus | 3 | Visual word recognition and semantic word processing (Stoeckel et al., 2009). |
| R caudal anterior cingulate gyrus* | 3 | Cognitive processes such as attention, salience and interference (Bush et al., 2000). |
| R pericalcarine cortex | 3 | Visual association cortex. |
| R precuneus* | 3 | Visuo-spatial imagery and episodic memory (see Supplementary Table 4). |
| R rostral middle frontal gyrus | 3 | Executive function; the region, as defined by Freesurfer, comprises the dorsolateral prefrontal cortex (Desikan et al., 2006; Kikinis et al., 2010), which is involved in working memory (Barbey et al., 2013) and attention and attention (Japee et al., 2015; Rosen et al., 1999). |
| R superior temporal gyrus | 3 | Visual information integration (Karnath, 2001; Shen et al., 2017). |
| L inferior parietal gyrus* | 2 | Association area at the temporo-parietal junction (Igelström and Graziano, 2017). |
| L accumbens area | 2 | Motor function and reward. |
| R thalamus | 2 | Relaying sensorimotor signals to the cortex. |
| R pallidum | 2 | Motor function and reward. |
| R posterior cingulate gyrus | 2 | Internally directed thought (Leech et al., 2011) and task management (Pearson et al., 2011). |
| R superior parietal gyrus* | 2 | See L superior parietal gyrus above. |
| L caudal anterior cingulate gyrus* | 1 | See R caudal anterior cingulate gyrus above. |
| L precuneus* | 1 | See R precuneus above. |
| R caudate | 1 | Goal-directed action. |
| R putamen | 1 | Movement regulation. |
| R hippocampus | 1 | Memory formation and spatial navigation (Eichenbaum, 2000). |
| R fusiform gyrus | 1 | Object and face recognition (Kleinhans et al., 2008; Pelphrey et al., 2007). |
| R inferior parietal gyrus* | 1 | See L inferior parietal gyrus above. |
| R inferior temporal gyrus | 1 | Visual processing and visual object recognition. |
| R isthmus of the cingulate gyrus | 1 | Connects the posterior cingulate cortex to the hippocampus and has a role in memory. |
| R middle temporal gyrus | 1 | Semantic memory processing. |
| R paracentral gyrus | 1 | Sensorimotor functions of the lower limb. |
| R pars triangularis | 1 | Semantic processing. |
| R superior frontal gyrus | 1 | Working memory and attention (Li et al., 2013), and contains the supplementary motor area. |

**Supplementary Table 6: Nodes in the processing speed subnetwork.** These are listed by the number of connections they make within this subnetwork, and associated function. * nodes which are present bilaterally in the subnetwork.

| | | | |
|---|---|---|---|
| ***Frontal*** | | ***Limbic*** | |
| CMFG | Caudal middle frontal gyrus | AC | Accumbens area |
| FP | Frontal pole | AM | Amygdala |
| LOFG | Lateral orbital frontal gyrus | CACG | Caudal anterior cingulate gyrus |
| MOFG | Medial orbital frontal gyrus | HI | Hippocampus |
| PaCG | Paracentral gyrus | ICG | Isthmus of the cingulate gyrus |
| POP | Pars opercularis | IN | Insula |
| POR | Pars orbitalis | PCG | Posterior cingulate gyrus |
| PTR | Pars triangularis | PHIG | Parahippocampal gyrus |
| PrCG | Precentral gyrus | RACG | Rostral anterior cingulate gyrus |
| RMFG | Rostral middle frontal gyrus | | |
| SFG | Superior frontal gyrus | | |
| ***Parietal*** | | ***Occipital*** | |
| IPG | Inferior temporal gyrus | CU | Cuneus cortex |
| PCU | Precuneus cortex | LG | Lingual gyrus |
| PoCG | Postcentral gyrus | LOG | Lateral occipital gyrus |
| SMG | Supramarginal gyrus | PCAL | Pericalcarine cortex |
| SPG | Superior parietal gyrus | | |
| ***Temporal*** | | ***Subcortical*** | |
| BSTS | Banks of the superior temporal sulcus | PA | Pallidum |
| EC | Entorhinal cortex | PU | Putamen |
| FG | Fusiform gyrus | TH | Thalamus |
| ITG | Inferior temporal gyrus | | |
| MTG | Middle temporal gyrus | CER | Cerebellum |
| STG | Superior temporal gyrus | | |
| TP | Temporal pole | | |
| TTG | Tranverse temporal gyrus | | |

**Supplementary Table 7: Node label abbreviations.**

### References

Barbey AK, Koenigs M, Grafman J. 2013. Dorsolateral prefrontal contributions to human working memory. *Cortex* **49**:1195–1205. doi:10.1016/j.cortex.2012.05.022

Bush G, Luu P, Posner MI. 2000. Cognitive and emotional influences in anterior cingulate cortex. *Trends Cogn Sci* **4**:215–222. doi:10.1016/S1364-6613(00)01483-2

Cavanna AE. 2007. The Precuneus and Consciousness. *CNS Spectr* **12**:545–552. doi:10.1017/S1092852900021295

Cavanna AE, Trimble MR. 2006. The precuneus: a review of its functional anatomy and behavioural correlates. *Brain* **129**:564–583. doi:10.1093/brain/awl004

Desikan RS, Ségonne F, Fischl B, Quinn BT, Dickerson BC, Blacker D, Buckner RL, Dale AM, Maguire RP, Hyman BT, Albert MS, Killiany RJ. 2006. An automated labeling system for subdividing the human cerebral cortex on MRI scans into gyral based regions of interest. *Neuroimage* **31**:968–980. doi:10.1016/j.neuroimage.2006.01.021

Eichenbaum H. 2000. A cortical–hippocampal system for declarative memory. *Nat Rev Neurosci* **1**:41–50. doi:10.1038/35036213

Grill-Spector K, Kushnir T, Edelman S, Avidan G, Itzchak Y, Malach R. 1999. Differential Processing of Objects under Various Viewing Conditions in the Human Lateral Occipital Complex. *Neuron* **24**:187–203. doi:10.1016/S0896-6273(00)80832-6

Igelström KM, Graziano MSA. 2017. The inferior parietal lobule and temporoparietal junction: A network perspective. *Neuropsychologia* **105**:70–83. doi:10.1016/j.neuropsychologia.2017.01.001

Japee S, Holiday K, Satyshur MD, Mukai I, Ungerleider LG. 2015. A role of right middle frontal gyrus in reorienting of attention: a case study. *Front Syst Neurosci* **9**. doi:10.3389/fnsys.2015.00023

Karnath H-O. 2001. New insights into the functions of the superior temporal cortex. *Nat Rev Neurosci* **2**:568–576. doi:10.1038/35086057

Kikinis Z, Fallon JH, Niznikiewicz M, Nestor P, Davidson C, Bobrow L, Pelavin PE, Fischl B, Yendiki A, McCarley RW, Kikinis R, Kubicki M, Shenton ME. 2010. Gray matter volume reduction in rostral middle frontal gyrus in patients with chronic schizophrenia. *Schizophr Res* **123**:153–159. doi:10.1016/j.schres.2010.07.027

Kleinhans NM, Richards T, Sterling L, Stegbauer KC, Mahurin R, Johnson LC, Greenson J, Dawson G, Aylward E. 2008. Abnormal functional connectivity in autism spectrum disorders during face processing. *Brain* **131**:1000–1012. doi:10.1093/brain/awm334

Leech R, Kamourieh S, Beckmann CF, Sharp DJ. 2011. Fractionating the Default Mode Network: Distinct Contributions of the Ventral and Dorsal Posterior Cingulate Cortex to Cognitive Control. *J Neurosci* **31**:3217–3224. doi:10.1523/JNEUROSCI.5626-10.2011

Li W, Qin W, Liu H, Fan L, Wang J, Jiang T, Yu C. 2013. Subregions of the human superior frontal gyrus and their connections. *Neuroimage* **78**:46–58. doi:10.1016/j.neuroimage.2013.04.011

Pearson JM, Heilbronner SR, Barack DL, Hayden BY, Platt ML. 2011. Posterior cingulate cortex: adapting behavior to a changing world. *Trends Cogn Sci* **15**:143–151. doi:10.1016/j.tics.2011.02.002

Pelphrey KA, Morris JP, McCarthy G, LaBar KS. 2007. Perception of dynamic changes in facial affect and identity in autism. *Soc Cogn Affect Neurosci* **2**:140–149. doi:10.1093/scan/nsm010

Rosen AC, Rao SM, Caffarra P, Scaglioni A, Bobholz JA, Woodley SJ, Hammeke TA, Cunningham JM, Prieto TE, Binder JR. 1999. Neural Basis of Endogenous and Exogenous Spatial Orienting: A Functional MRI Study. *J Cogn Neurosci* **11**:135–152. doi:10.1162/089892999563283

Shen W, Yuan Y, Liu C, Luo J. 2017. The roles of the temporal lobe in creative insight: an integrated review. *Think Reason* **23**:321–375. doi:10.1080/13546783.2017.1308885

Shultz S, Lee SM, Pelphrey K, McCarthy G. 2011. The posterior superior temporal sulcus is sensitive to the outcome of human and non-human goal-directed actions. *Soc Cogn Affect Neurosci* **6**:602–611. doi:10.1093/scan/nsq087

Stoeckel C, Gough PM, Watkins KE, Devlin JT. 2009. Supramarginal gyrus involvement in visual word recognition. *Cortex* **45**:1091–1096. doi:10.1016/j.cortex.2008.12.004

Uddin LQ, Nomi JS, Hébert-Seropian B, Ghaziri J, Boucher O. 2017. Structure and Function of the Human Insula. *J Clin Neurophysiol* **34**:300–306. doi:10.1097/WNP.0000000000000377

Zalesky A, Cocchi L, Fornito A, Murray MM, Bullmore E. 2012. Connectivity differences in brain networks. *Neuroimage* **60**:1055–1062. doi:10.1016/j.neuroimage.2012.01.068

Zalesky A, Fornito A, Harding IH, Cocchi L, Yücel M, Pantelis C, Bullmore ET. 2010. Whole-brain anatomical networks: Does the choice of nodes matter? *Neuroimage* **50**:970–983. doi:10.1016/j.neuroimage.2009.12.027
